# Selective Modulation of Evidence Accumulation by Hippocampal Theta Oscillations during Mnemonic Decision-Making

**DOI:** 10.1101/2025.07.18.25331749

**Authors:** Pei L. Robins, Jessica R. Gilbert, Bruce Luber, Nadia Mustafa, Eesha Bharti, Jeffrey D. Stout, Frederick W. Carver, Zhi-De Deng

## Abstract

A core function of episodic memory is to distinguish between overlapping experiences by converting similar inputs into distinct, non-overlapping representations, a process termed pattern separation. While anatomical models emphasize the role of specific hippocampal subfields, particularly the dentate gyrus, CA3, and CA1, less is known about how these computations unfold over time and influence memory-based decisions. Here, we use source-localized magnetoencephalography and computational modeling to examine how theta oscillations from the hippocampus as a whole are related to evidence accumulation during mnemonic discrimination. Participants performed the Mnemonic Similarity Task, in which they classified Repeat, Lure, and Foil images as “Old,” “Similar,” or “New.” Event-related spectral and source activity confirmed reliable hippocampal engagement during the task despite its anatomical depth. We fit a hierarchical Linear Ballistic Accumulator model to behavioral data, estimating trial-by-trial drift rates as a latent index of mnemonic evidence accumulation, and examined whether hippocampal theta power predicted these dynamics. Left hippocampal theta was negatively associated with drift toward “New” responses on lure trials, while right hippocampal theta was positively associated with drift toward “Similar” responses on foil trials. These effects suggest that hippocampal theta selectively indexes partial-match sensitivity, with consequences that are beneficial or costly depending on whether the stimulus has a encoded memory counterpart. However, direct comparisons between hemispheres did not yield credible differences. These findings offer preliminary evidence that trial-level hippocampal theta fluctuations are related to the dynamics of memory-guided decision-making, and demonstrate the feasibility of linking deep-source MEG recordings to computational models of evidence accumulation.

## 1 INTRODUCTION

Episodic memory enables the recollection of specific past experiences not only by preserving their content, but also by differentiating among events that overlap in time, space, or perceptual detail (Eichenbaum, 2017, Tulving, 1983). This ability to disambiguate similar experiences is thought to rely on a process known as pattern separation, in which inputs such as events that share temporal, spatial, or perceptual features, are transformed into orthogonalized neural representations to minimize interference (O’Reilly & McClelland, 1994, Yassa & Stark, 2011). The transformation is implemented through complementary mechanisms within the hippocampal circuit, in particular, the dentate gyrus (DG), and the CA3 and CA1 subfields. The DG provides sparse, non-overlapping representations of cortical inputs through low activation rates and random connectivity patterns, which decorrelate incoming signals (Leutgeb et al., 2007, Rolls, 2013, Treves & Rolls, 1994, Yassa & Stark, 2011). In contrast, the CA3 subfield, with its dense recurrent collaterals, supports autoassociative storage and pattern completion by enabling retrieval from partial cues. CA3 can also contribute to pattern separation under conditions of sufficiently distinct input or strong DG drive, exhibiting flexible dynamics depending on input similarity (Leutgeb et al., 2007, Yassa & Stark, 2011). Recent physiological work further supports this computational architecture: Allegra et al. demonstrated that while the DG consistently decorrelates similar inputs regardless of behavioral outcome, CA1 selectively reflects behaviorally relevant distinctions, suggesting that downstream hippocampal regions gate and interpret the output of DG–CA3 computations according to task demands (Allegra et al., 2020). These complementary properties allow the DG–CA3–CA1 circuit to flexibly support both memory discrimination and generalization. Increasingly, research suggests that this balance is not only anatomically distributed but also temporally orchestrated. In particular, hippocampal theta oscillations (4–8 Hz) have emerged as a dynamic scaffold for coordinating encoding and retrieval processes.

These oscillations organize neural activity into alternating phases that bias the hippocampus toward either external input or internal retrieval: during one phase of the theta cycle, entorhinal input dominates, supporting pattern separation and encoding; during the opposite phase, CA3 input prevails, promoting pattern completion and retrieval (Colgin, 2013, Hasselmo et al., 2002) Recent findings in humans provide direct empirical support for this theta-gamma phase opposition, demonstrating that hippocampal gamma amplitude is coupled to the theta peak during encoding and the theta trough during retrieval, with the degree of this opposition predicting later memory performance (Saint Amour di Chanaz et al., 2023). Moreover, volitional learning increases hippocampal theta power and promotes a sophisticated phase coding regime, in which the semantic similarity of items determines their intracycle reactivation timing, effectively segregating conceptually similar items into more distant theta phases (Pacheco Estefan et al., 2021). This functional segregation minimizes interference between incoming and stored information, and evidence from intracranial electroencephalography (iEEG) confirms that increased theta power preceding stimulus onset predicts later memory success, highlighting theta’s role in establishing a preparatory mnemonic state (Fell et al., 2011, Guderian et al., 2009). Buzsáki further conceptualizes theta as a temporal organizer of hippocampal assemblies, aligning neuronal firing with windows of synaptic plasticity (Buzsáki, 2002). Theta’s role extends to retrieval: increased oscillatory power and inter-regional synchronization facilitate the reinstatement of previously encoded representations (Osipova et al., 2006). In behavioral contexts requiring memory-guided decisions, the theta rhythm helps resolve competition among overlapping traces by structuring neural computation into discrete, state-specific epochs (Hasselmo, 2005, Herweg et al., 2020).

Despite this compelling framework, much of the empirical literature relies on trial-averaged estimates of theta power. This approach leaves unresolved whether trial-to-trial fluctuations in hippocampal theta contribute to memory performance and decision-making in contexts demanding mnemonic discrimination. We hypothesized that if theta orchestrates the encoding–retrieval balance critical for pattern separation, then moment-to-moment fluctuations in theta power should predict the quality of evidence accumulation during mnemonic decisions. While other neural oscillations support decision-making more generally (e.g., beta in sensory evidence accumulation and cognitive set maintenance (Engel & Fries, 2010); alpha in attentional gating and inhibition of task-irrelevant information (Klimesch, 2012)), hippocampal theta is uniquely tied to the mechanistic segregation of encoding and retrieval cycles. Theta, thus, provides the most theoretically motivated frequency band for linking hippocampal computation to evidence accumulation during pattern separation.

The Mnemonic Similarity Task (MST) provides a robust and well-validated tool for studying the neural mechanisms of mnemonic discrimination (Stark et al., 2019). The task consists of two phases: during encoding, participants view a series of images depicting common everyday objects; during retrieval, they are asked to classify each item as “old,” “similar,” or “new” from a mixed set that includes repeated items, similar but non-identical lures, and novel items. Crucially, accurate identification of lures as “similar” is interpreted as behavioral evidence of pattern separation, while misclassification of lures as “old” is often taken to reflect pattern completion or memory generalization. This interpretive framework maps onto the proposed computational architecture of the hippocampus described above: activity in the DG is thought to support successful lure discrimination, whereas the CA3 subfield modulates the tradeoff between separation and completion based on input similarity (Bakker et al., 2008, Berron et al., 2016, Yassa et al., 2011). Functional magnetic resonance imaging (fMRI) studies have consistently implicated DG/CA3 activity in lure discrimination, yet the slow temporal resolution of fMRI limits access to the rapid computations that shape memory decisions. Moreover, MST decisions are inherently ambiguous: lures are designed to resemble repeats, and accurate responding requires resolving this perceptual and mnemonic overlap in real time. For this reason, the MST offers a valuable context not only for probing hippocampal function but also for examining how memory signals are transformed into decisions under uncertainty. However, the task’s behavioral outcome—response time (RT)—reflects the aggregate duration of multiple cognitive processes, including visual processing, evidence accumulation, and motor execution (button press). As a result, it is difficult to isolate specific decision-related computations. To access the quality and dynamics of mnemonic evidence more directly, computational modeling is required.

Sequential sampling models such as the Linear Ballistic Accumulator (LBA) offer a principled framework for disentangling the cognitive processes underlying memory-based decisions (Brown & Heathcote, 2008, Donkin, Brown, Heathcote, & Wagenmakers, 2011). The model decomposes observed choices and RTs into latent parameters, among which *drift rate* quantifies the quality and speed of evidence accumulation. This parameter is particularly well-suited to capture trial-level variability in mnemonic strength, especially in tasks like the MST that demand fine-grained discrimination under uncertainty. Demonstrating this application, Banavar et al. applied the LBA to MST performance and identified drift rate as a meaningful and interpretable marker of mnemonic evidence strength (Banavar et al., 2024). Electrophysiological correlates have been observed, as studies have demonstrated that both event-related potentials and oscillatory dynamics track continuous variation in decision evidence, across both perceptual and mnemonic domains, and correlate with model-derived measures of decision processing (Addante et al., 2011, Philiastides & Sajda, 2006, Ratcliff et al., 2009). In parallel, neuroimaging studies have identified drift rate correlates in parietal and prefrontal cortices during memory-based decisions (Mueller et al., 2017, Mulder et al., 2014), reinforcing the broader utility of these models for linking cognitive computations to brain activity.

Despite these advances, no studies to date have directly linked measures of single-trial neural dynamics, such as hippocampal theta oscillations, to computational parameters of decision-making during mnemonic discrimination. This lack of integration leaves a critical gap in our understanding: while computational models offer mechanistic insight into behavior, and neural signals reflect ongoing cognitive processing, the two have yet to be combined at the level of individual memory decisions. Bridging this divide is essential for connecting cognitive theory to neural mechanisms in a temporally precise and functionally interpretable manner.

In the present study, we address this gap by combining source-localized magnetoencephalography (MEG) with LBA modeling during the MST. Our primary aim was to determine whether trial-by-trial fluctuations in hippocampal theta power predict the rate of evidence accumulation during memory decisions, and whether these relationships differ across hemispheres. Although the hippocampus has been widely implicated in pattern separation, many prior studies have either pooled across hemispheres, focused on long-axis (anterior–posterior) gradients, or examined subfield-level effects without reporting lateralization. Yet, several lines of research suggest that the left and right hemispheres may support distinct contributions to memory. For example, the Hemispheric Encoding/Retrieval Asymmetry (HERA) model (Tulving et al., 1994) proposed a functional division between left-lateralized encoding and right-lateralized retrieval processes in episodic memory. While HERA emphasized prefrontal contributions, more recent studies point to anatomical, functional, and oscillatory asymmetries across the left and right hippocampus, suggesting that lateralized dynamics may extend beyond the cortex (Penner et al., 2022, Stevenson et al., 2020). Building on this literature, our MEG source modeling approach allowed us to estimate hippocampal activity separately in each hemisphere and to study lateralization in theta-related decision dynamics. Prior MEG work has demonstrated that hippocampal responses can be detected under recognition memory conditions when appropriate source modeling techniques are used, including beamforming and spectral analyses in the theta band (Riggs et al., 2009). By linking oscillatory neural dynamics directly to computational indices of decision-making, our approach offers a novel framework for identifying the mechanistic role of hippocampal theta in evidence accumulation. In doing so, it advances the integration of cognitive and neural accounts of pattern separation in human episodic memory.

## 2 METHODS

### 2.1 Participants

Twenty healthy volunteers were initially enrolled in the study, which was conducted under a National Institutes of Health (NIH) technical development protocol aimed at advancing neuroimaging methods for studying mood and anxiety disorders (ClinicalTrials.gov ID: NCT00397111). Six participants were excluded from the analysis due to missing structural magnetic resonance imaging (MRI) scans (*n* = 2), poor head localization (*n* = 1), or poor behavioral task performance (*n* = 3), the latter due to drowsiness or misunderstanding task instructions. The final sample consisted of fourteen participants (13 female, 1 male), with a mean age of 32 years (range: 21–54). We acknowledge that the predominantly female composition limits generalizability, as sex differences have been documented in hippocampal function, episodic memory, and pattern separation performance (Herlitz et al., 1997, Yagi et al., 2016). All participants were right-handed, reported normal or corrected-to-normal vision, and had no history of neurological or psychiatric disorders. Written informed consent was obtained from all participants, and all procedures were approved by the Institutional Review Board of the National Institute of Mental Health (NIMH).

### 2.2 Behavioral Task

Participants performed the MST (Stark et al., 2019) while seated in the MEG scanner (MEG Core Facility, NIH). The task, designed specifically to probe hippocampal pattern separation, comprised an encoding phase and a retrieval phase, implemented using PsychoPy2 (Peirce et al., 2019). During the encoding phase, participants viewed 120 images of everyday objects, making “Indoor”/”Outdoor” judgments for each item. During the retrieval phase, participants classified 360 images as “Old,” “Similar,” or “New.” These images included 120 previously viewed items (Repeats), 120 similar but non-identical items (Lures), and 120 novel items (Foils). All stimuli were presented for 2 seconds, followed by a 0.5-second interstimulus interval between the subsequent trials (Figure 1). The retrieval phase was intentionally extended to 360 trials (compared with 192 trials in prior studies) to enable sufficient signal-to-noise ratio for reliable estimation of evoked fields and deep hippocampal source activity using MEG. Although this increase in trial count necessarily elevates mnemonic load and proactive interference, it was required to support the neurophysiological aims of the study.

**FIGURE 1.**
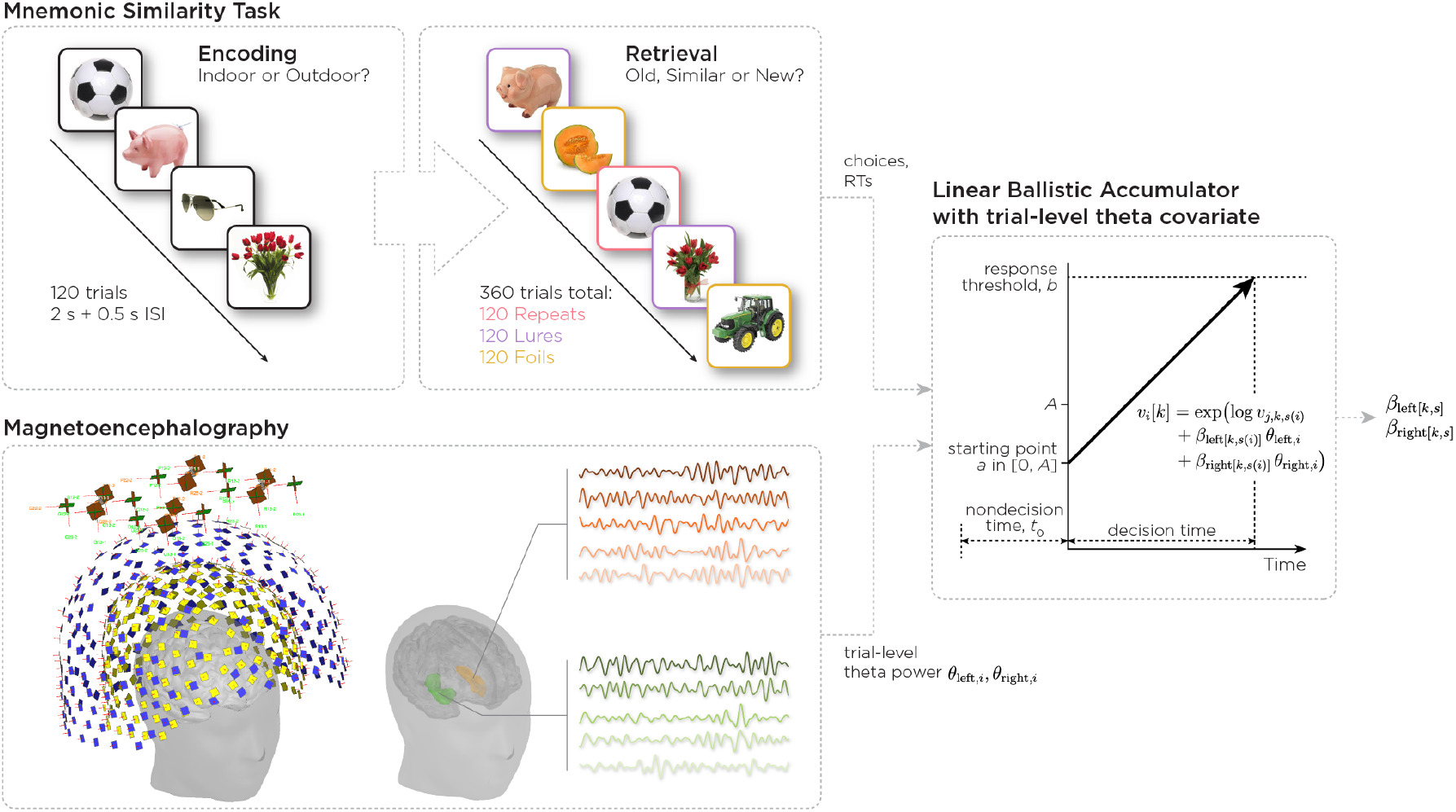
Overview of experimental design and modeling approach. Participants performed a two-phase Mnemonic Similarity Task during magnetoencephalography. During encoding, participants viewed 120 common objects and made an “indoor”/”outdoor” judgment for each item. At retrieval, participants viewed 360 images comprising three stimulus types in equal proportions: Repeats, Lures, and Foils. For each item, participants made a three-alternative forced choice response: “Old”, “Similar”, or “New”. Magnetoencephalography signals were recorded continuously, and hippocampal time series were reconstructed using source localization techniques. Trial-level theta-band power (4–8 Hz) was extracted separately for the left and right hippocampus. We fit a hierarchical Linear Ballistic Accumulator (LBA) model to the joint distribution of choices and response times. The LBA conceptualizes decisions as a race between accumulators, with evidence accumulating linearly for each response option at rate *v* (drift rate) from random starting points uniformly distributed in [0, *A*] until one accumulator reaches threshold *b*. The total response time comprises decision time plus non-decision time *t*_0_. Critically, drift rates were modulated by trial-level theta power from both hemispheres via regression coefficients *β*_left [*k,s*]_ and *β*_right[*k,s*]_ for each response option *k* and stimulus type *s*, allowing us to test how lateralized hippocampal oscillations influence evidence accumulation for different memory judgments.

### 2.3 Behavioral Indices: Lure Discrimination Index and Recognition Score

To quantify memory performance during the MST, we computed two behavioral indices: the *Lure Discrimination Index (LDI)* and the *Recognition Score (REC)*. The LDI reflects the ability to distinguish similar lures from novel foils, adjusted for response bias, and is defined as:

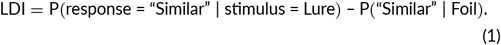

The Recognition Score (REC) reflects corrected recognition memory by adjusting for false alarms to novel stimuli. It is defined as:

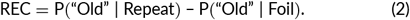

These definitions follow prior work characterizing behavioral performance in the MST (Stark et al., 2019).

### 2.4 Imaging Data Acquisition and Preprocessing

Structural MRI scans were acquired using a 3T GE (General Electric) gradient-echo scanner equipped with a 32-channel head coil. A high-resolution T1-weighted MP-RAGE sequence was used to obtain anatomical images for source modeling [field of view = 256 mm^2^; voxel size = 1 mm isotropic; echo time (TE) = minimum full; inversion time (TI) = 1100 ms; flip angle = 7°; bandwidth = 25 Hz/pixel].

To measure neural activity during the task, we recorded MEG data using a whole-head CTF MEG system (CTF Systems, Inc., Canada), which includes 275 radial first-order gradiometer SQUID channels. Head position was continuously monitored using three localization coils placed at fiducial landmarks. These coils were positioned 1.5 cm above the nasion, and at left and right preauricular points (1.5 cm anterior to each tragus along a line extending through the outer canthus of the eye). Fiducial locations were co-registered to each participant’s MRI using Brainstorm (Tadel et al., 2011). Data were acquired continuously with an online synthetic third-order gradiometer configuration to suppress environmental magnetic interference. Data were primarily sampled at 600 Hz with an acquisition bandwidth of 0–150 Hz. Due to variability in hardware settings, recordings of three participants were sampled at 1200 Hz.

Following the acquisition, all data analysis was performed in Brainstorm. Data sampled at 1200 Hz were downsampled to 600 Hz, and all data were bandpass filtered between 1 and 100 Hz. A 60 Hz notch filter was applied to suppress line noise. Artifact correction was performed on the continuous data using Infomax independent component analysis (ICA) to identify and remove components associated with ocular, cardiac, and muscle artifacts. Components were manually evaluated using a combination of time course, topography, and frequency characteristics. On average, 4.4 (range 3–8) out of 20 ICA components were removed for each participant. Following ICA, the data were segmented into epochs ranging from 200 ms before to 1000 ms after stimulus onset, and trials containing residual high-amplitude transients were excluded from further analysis.

Trials with RTs less than 200 ms or no response were also excluded from the analysis, as such fast responses are unlikely to reflect genuine stimulus-driven decisions. Following preprocessing, 4,907 trials were retained across 14 participants (mean: 351, range: 314–359).

### 2.5 Source Modeling and Event-Related Signal Extraction

Individualized head models were computed using the overlapping spheres method, and source estimation was performed using a depth-weighted dynamic statistical parametric mapping (dSPM) approach (Dale et al., 2000). The noise covariance matrix was estimated using the raw MEG data using the 200-ms pre-stimulus baseline. Dipole orientations were constrained normal to the cortical and hippocampal surfaces (Attal et al., 2009, Attal & Schwartz, 2013), as defined by anatomical parcellation from FreeSurfer 7.4.1 Automatic Subcortical Segmentation (ASEG) atlas (Fischl, 2013).

Source time series were extracted from dSPM values within the left and right hippocampal regions of interest (ROIs). To account for ambiguity in dipole polarity and to derive a representative signal per hemisphere, we computed the root-mean-square (RMS) across all dipoles within each ROI. These RMS time series were baseline-corrected using a 200-ms pre-stimulus window and transformed into *Z*-scores, yielding standardized, stimulus-locked estimates of hippocampal activity.

To verify that hippocampal sources exhibited task-evoked responses with sufficient signal-to-noise ratios, we computed event-related fields (ERFs) by averaging the source-localized RMS time series across trials for each task phase and stimulus condition. Visual inspection of group-averaged ERFs focused on response magnitude, latency, and temporal dynamics, confirming robust and temporally consistent hippocampal activation in response to MST stimuli.

### 2.6 Event-Related Spectral Perturbation Analysis

To characterize stimulus-locked changes in hippocampal oscillatory activity across a broad frequency range (from 1–100 Hz), we computed event-related spectral perturbation (ERSP) maps from source-localized hippocampal time series. Time–frequency decomposition was performed separately for each trial using complex Morlet wavelets. Spectral power was obtained by squaring the magnitude of the complex wavelet coefficients. To normalize power values and emphasize task-related changes, ERSPs were baseline-corrected using a 200-ms pre-stimulus interval. Power at each time–frequency point was expressed as percent change relative to the mean baseline power:

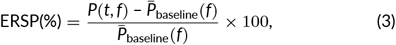

where *P*(*t, f*) denotes power at time *t* and frequency *f*, and 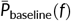 is the mean power during the baseline period at that frequency. ERSP maps were averaged across trial conditions separately for the left and right hippocampus during the encoding and retrieval phases to verify robust theta-band engagement during the task.

### 2.7 Single-Trial Theta Power Estimation

To quantify trial-by-trial fluctuations in hippocampal oscillatory activity, we estimated theta-band (4–8 Hz) power from source-localized time series in the left and right hippocampus during the encoding and retrieval phase. The dSPM time series was bandpass filtered from 4–8 Hz using a zero-phase finite impulse response filter, and the analytic signal was computed via the Hilbert transform. Instantaneous power was obtained by squaring the magnitude of the resulting complex signal and averaging over the 1000-ms post-stimulus window. The 0–1000 ms post-stimulus window was selected on the basis that stimulus-evoked hippocampal activity was negligible beyond 1000-ms in our ERF results, consistent with prior work demonstrating that EEG activity during recognition memory is concentrated within this interval (Anderson et al., 2017). This window was further supported by the observed mean response times, which ranged from approximately 938 to 1085-ms across stimulus–response conditions; since decision-related neural activity necessarily precedes the motor response, the neural correlates of evidence accumulation are expected to fall within this interval on the majority of trials. The power values were then normalized using percentage change from the 200-ms pre-stimulus baseline, providing baseline-corrected estimates of stimulus-evoked theta activity on a trial-by-trial basis.

### 2.8 Subsequent Memory Effect Analysis

To examine whether encoding-phase hippocampal theta power predicted later retrieval success, we performed Subsequent Memory Effect (SME) analyses. For each participant, we linked encoding trials to their corresponding retrieval outcomes. Trials were categorized as Hits (items correctly recognized as “Old”); or Misses (items incorrectly judged as “Similar” or “New”). We computed subject-level mean theta power for Hits and Misses separately for each hemisphere. Group-level differences were assessed using paired *t*-tests.

### 2.9 Linear Ballistic Accumulator Model

We fit a hierarchical Bayesian LBA model (Brown & Heathcote, 2008) to jointly model choice and RT data. The LBA represents each response option as an independent evidence accumulator that races toward a common decision threshold. On each trial, the first accumulator to reach threshold determines the response and the decision time; the observed RT equals the decision time plus a non-decision component. The model included five parameters: drift rate (*v*) governs the mean speed of evidence accumulation for each response option; starting-point variability (*A*) captures trial-to-trial variability in initial evidence; threshold (*b*) specifies the amount of evidence required to respond; non-decision time (*t*_0_) accounts for visual processing and motor execution; drift-rate variability (*sv*) captures across-trial variability in drift around its mean.

#### 2.9.1 Hierarchical Structure

We implemented a hierarchical model with parameters indexed by participant (*j* = 1 … *J*), response option (*k* = 1 … 3: “New”, “Similar”, “Old”), and stimulus type (*s* = 1 … 3: Foil, Lure, Repeat). Drift rates varied by participant, response option, and stimulus type. In contrast, *A, b*, and *t*_0_ varied by participant only. To ensure positivity, we estimated all positive parameters on the log scale and parameterized the threshold as

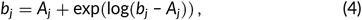

which enforces *b*_*j*_ > *A*_*j*_.

We used a non-centered parameterization for hierarchical effects:

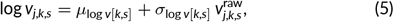

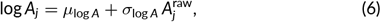

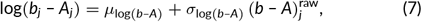

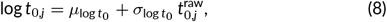

where *µ* terms denote group-level means, *σ* terms denote group-level standard deviations, and raw deviations were assigned standard Normal priors.

### 2.9.2 Theta Modulation of Drift Rates

To test whether hippocampal theta modulates evidence accumulation, we modeled trial-by-trial hippocampal theta power as a predictor of drift rate. For trial *i*, the drift rate for response option *k* was

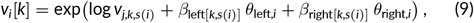

where *θ*_left,*i*_ and *θ*_right,*i*_ are left and right hippocampal theta power on trial *i, z*-scored across all trials in the analysis set (separately for each hemisphere), and *β*_left[*k,s*]_ and *β*_right[*k,s*]_ are regression coefficients that vary by response option and stimulus type. Positive values of *β* indicate that higher theta power increases drift rate (stronger evidence accumulation) for that response–stimulus combination. Because *θ* is standardized, *β* reflects the change in log drift rate associated with one standard deviation increase in theta power.

#### 2.9.3 Priors

We specified weakly informative priors centered on plausible values from prior LBA applications in recognition memory (Donkin, Brown, & Heathcote, 2011) (Table 1). Group-level means were assigned Normal priors with standard deviation 0.5 on the log scale. For example, *µ*_log *v*_ ∼ 𝒩 (log(1.5), 0.5) places substantial mass on drift rates commonly observed in multi-choice paradigms. Similarly, 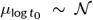 (log(0.2), 0.5) centers non-decision time at 200 ms while permitting a broad range of values.

**TABLE 1.**
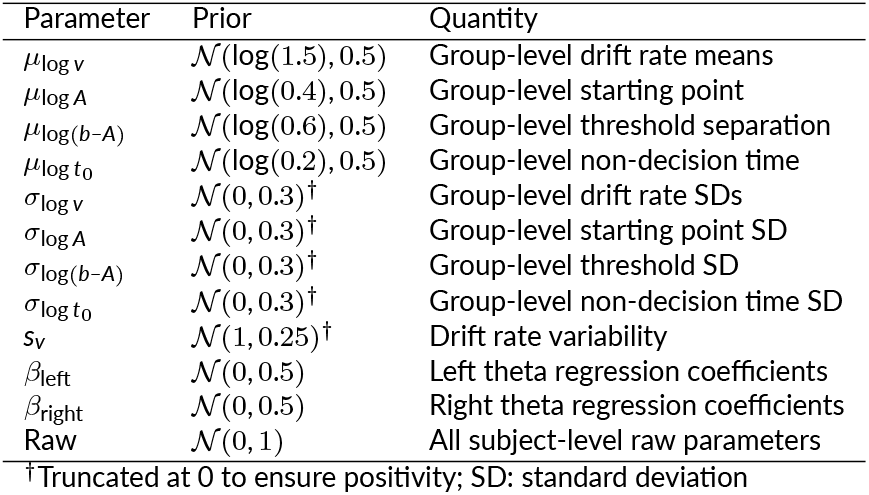
Prior distributions for model parameters.

| Parameter | Prior | Quantity |
| --- | --- | --- |
| $\mu_{\log v}$ | $\mathcal{N}(\log(1.5), 0.5)$ | Group-level drift rate means |
| $\mu_{\log A}$ | $\mathcal{N}(\log(0.4), 0.5)$ | Group-level starting point |
| $\mu_{\log(b-A)}$ | $\mathcal{N}(\log(0.6), 0.5)$ | Group-level threshold separation |
| $\mu_{\log t_0}$ | $\mathcal{N}(\log(0.2), 0.5)$ | Group-level non-decision time |
| $\sigma_{\log v}$ | $\mathcal{N}(0, 0.3)^\dagger$ | Group-level drift rate SDs |
| $\sigma_{\log A}$ | $\mathcal{N}(0, 0.3)^\dagger$ | Group-level starting point SD |
| $\sigma_{\log(b-A)}$ | $\mathcal{N}(0, 0.3)^\dagger$ | Group-level threshold SD |
| $\sigma_{\log t_0}$ | $\mathcal{N}(0, 0.3)^\dagger$ | Group-level non-decision time SD |
| $sv$ | $\mathcal{N}(1, 0.25)^\dagger$ | Drift rate variability |
| $\beta_{\text{left}}$ | $\mathcal{N}(0, 0.5)$ | Left theta regression coefficients |
| $\beta_{\text{right}}$ | $\mathcal{N}(0, 0.5)$ | Right theta regression coefficients |
| Raw | $\mathcal{N}(0, 1)$ | All subject-level raw parameters |

<sup>†</sup>Truncated at 0 to ensure positivity; SD: standard deviation

Group-level standard deviations were assigned 𝒩 (0, 0.3)^†^ priors to mildly regularize between-subject variability while preserving flexibility. Drift-rate variability was assigned *sv* ∼ 𝒩 (1, 0.25)^†^. The theta regression coefficients were assigned 𝒩 (0, 0.5) priors; because theta power was *z*-scored, a coefficient of 0.5 corresponds to a multiplicative change in drift of exp(0.5) ≈ 1.65 per standard deviation of theta. Subject-level raw deviations were assigned 𝒩 (0, 1) priors, consistent with non-centered hierarchical parameterizations (Betancourt & Girolami, 2015).

### 2.9.4 Model Estimation

We fit the model using Hamiltonian Monte Carlo (HMC) with the No-U-Turn Sampler (NUTS) as implemented in RStan (Stan Development Team, 2024). We ran four chains for 5,000 iterations each, discarding the first 1,000 iterations as warmup, for a total of 16,000 post-warmup draws. We set adapt_delta to 0.95 and the maximum tree depth to 12 to reduce divergent transitions in this high-dimensional posterior. We initialized chains using conservative values informed by the observed RT distribution. Specifically, we initialized *t*_0_ at 40% of the minimum observed RT to ensure *t*_0_ remained below all observed response times.

### 2.10 Model Evaluation

Model convergence was assessed using the potential scale reduction factor, 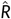 (Gelman & Rubin, 1992), along with bulk and tail effective sample sizes and visual inspection of trace plots. We also recorded the percentage of post-warmup divergent transitions and treated values < 0.1 as indicating acceptable posterior geometry.

We evaluated model adequacy using posterior predictive checks (PPCs). For each of 500 draws from the joint posterior, we simulated a full dataset matching the observed design (subjects, trials, stimulus types, response options, and observed trial-level theta covariates). On each simulated trial, we computed drift rates using Equation 9. We sampled start points uniformly from [0, *A*_*j*_] and generated trial-wise drift rates by perturbing the mean drift with Normal noise of standard deviation *s*_*v*_. The fastest accumulator determined the simulated choice and decision time, and we obtained simulated RTs by adding the corresponding *t*_0,*j*_. We summarized PPCs using three graphical checks. First, we overlaid observed RT densities with simulated RT densities, separately for each response option and stimulus type. Second, we compared observed RT quantiles (10th, 30th, 50th, 70th, 90th percentiles) with posterior predictive intervals for those quantiles, following established practice in the LBA literature (Brown & Heathcote, 2008, Donkin, Brown, Heathcote, & Wagenmakers, 2011). Third, we compared observed choice proportions with posterior predictive intervals for choice proportions within each stimulus type. Together, these checks indicated that the model reproduced both RT distributions and choice patterns across stimulus–response combinations.

Out-of-sample predictive accuracy was assessed using leave-one-out cross-validation (LOO-CV) using the loo package in R (Vehtari et al., 2017). We used Pareto *k* diagnostics to identify observations with undue influence on the LOO estimate, with *k* < 0.7 as indicating reliable estimates.

### 2.11 Alternative models

To test whether the theta–behavior relationship was specific to evidence accumulation, we fit two alternative LBA variants in which trial-level theta modulated either boundary separation (*b*) or starting-point range (*A*) (see Supplementary Materials for detailed methods). LOO-CV indicated that predictive accuracy was statistically indistinguishable across the three models (Supplementary Table S3). However, only the drift-rate model recovered a condition-specific theta signal; in the threshold and starting-point models, regression coefficients were uniformly small and their credible intervals spanned zero (Supplementary Figure S4). This pattern is consistent with theta being expressed preferentially through evidence accumulation rather than through boundary setting or starting-point variability.

## 3 RESULTS

### 3.1 Behavioral Performance and Drift Rate Estimates

To quantify behavioral performance during the MST, we computed two accuracy-based indices that separately captured recognition and discrimination. The mean REC was 0.57, indicating a moderate ability to distinguish previously seen items from novel foils; however, the mean REC was lower than typically observed in the standard MST protocol. To assess potential time-on-task effects, we examined recognition performance separately for the first and second halves of the retrieval phase. Descriptively, REC was higher in the first half (REC ≈ 0.6) than in the second half (REC ≈ 0.5), suggesting a decline in recognition accuracy during the extended task portion, although this difference did not reach statistical significance. The mean LDI was 0.33, consistent with normative values observed in large samples of healthy adults (Banavar et al., 2024).

Next, we examined the proportion of responses (Figure 2A) and the mean RTs across stimulus–response combinations (Figure 2B). Behavioral performance showed a low proportion of “Similar” responses relative to “Old” or “New” responses, similar to the proportion of responses observed in a large sample of healthy adults (Banavar et al., 2024). Trials in which participants responded “Similar” were generally observed with longer RTs than “Old” or “New” responses, consistent with the increased difficulty of correctly rejecting lures. However, because RTs reflect the aggregate of multiple cognitive operations, they offer limited specificity about the decision process itself. To more directly infer the quality and dynamics of mnemonic evidence, we applied the LBA model to estimate the underlying rate of evidence accumulation for each response type. Drift rates exhibited a strong diagonal structure (Figure 2C), with the fastest accumulation for stimulus-congruent responses (diagonal cells: “Old”|Repeat, “Similar”|Lure, “New”|Foil). In contrast, the most stimulus-incongruent pairings (“Old”|Foil; “New”|Repeat) showed the lowest drift rates, consistent with weak evidence for these accumulators.

**FIGURE 2.**
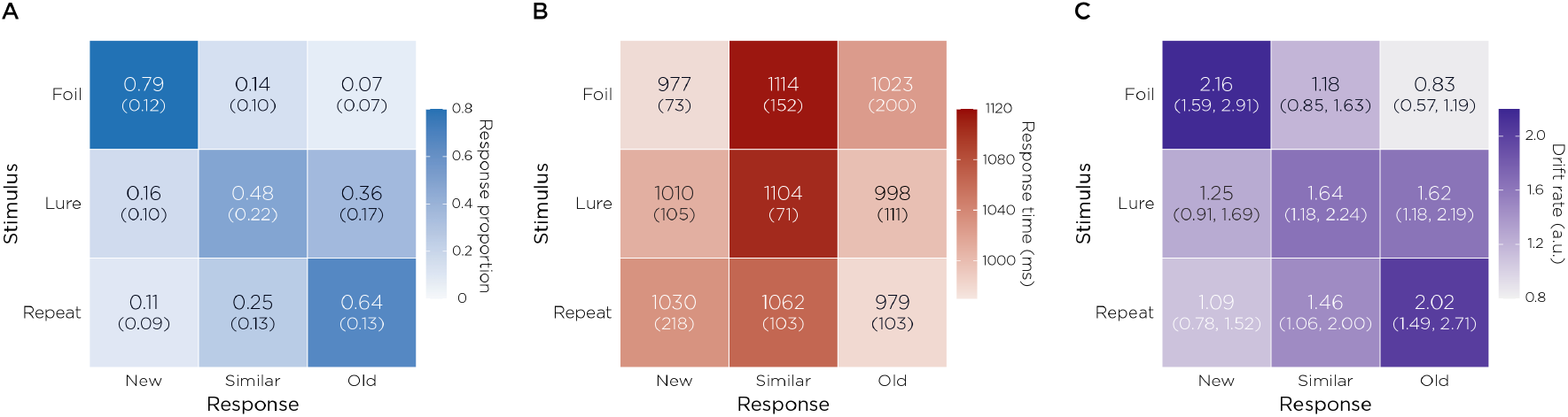
Behavioral performance and model-estimated drift rates. (A) Response proportions for each stimulus–response combination. Values represent mean proportions across participants, with standard deviations in parentheses. Participants showed high accuracy for Repeats (64% “Old” responses) and Foils (79% “New” responses), with moderate performance on Lures (48% “Similar” responses). (B) Mean response times (ms, standard deviations in parentheses) for each stimulus–response combination. Correct responses (diagonal) were generally faster than errors. (C) Posterior mean drift rates (*v*, in arbitrary units, with 95% credible intervals in parentheses) estimated by the hierarchical LBA model for each stimulus–response combination, averaged across participants. Higher drift rates indicate stronger evidence accumulation for that response option. The model captured key patterns in the data: highest drift rates along the diagonal (correct responses).

### 3.2 Hippocampal Event-Related Fields and Event-Related Spectral Perturbation

To establish whether hippocampal sources were reliably engaged during the task, we examined ERFs derived from source-localized time series (Figure 3). Stimulus-locked responses were evident in both hemispheres.

**FIGURE 3.**
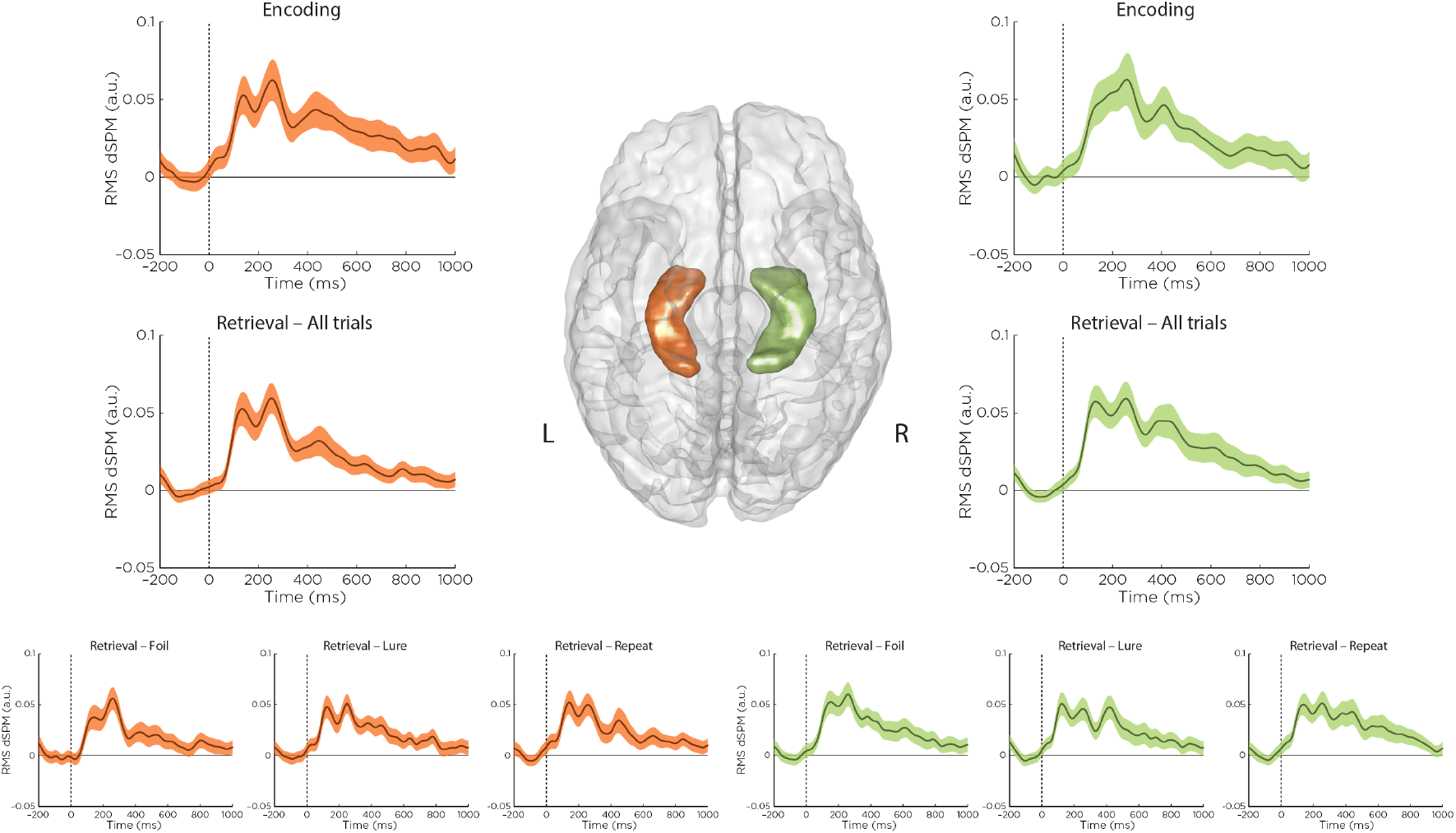
Average source-localized time series were extracted from left (orange) and right (green) hippocampi, computed as root-mean-square (RMS) of dynamic statistical parametric mapping (dSPM) *Z*-score values, and baseline-corrected using a 200-ms pre-stimulus window. Top row: Event-related fields (ERFs) during the encoding phase of the Mnemonic Similarity Task; Middle row: ERFs during the retrieval phase, averaged across all stimulus types; Bottom row: Retrieval-phase ERFs separated by stimulus type (repeats, lures, foils). Although dSPM values are shown in arbitrary units (a.u.) due to *Z*-scoring prior to averaging, stimulus-evoked responses were observed in both hemispheres. Narrow standard error bands indicate high consistency across participants.

Across both task phases, the group-averaged time courses revealed a well-defined response starting at approximately 100 ms after stimulus onset, followed by a polyphasic wave peaking between 200–300 ms, and a smaller subsequent peak between 400–500 ms. Importantly, the narrow standard error bands indicated consistent temporal dynamics across participants. Retrieval-phase responses were broadly similar across stimulus conditions (Repeat, Lure, Foil), with no strong qualitative differences between the left and right hippocampi. Together, these results demonstrate reliable stimulus-evoked hippocampal activity and provide a foundation for analyzing trial-level theta fluctuations in relation to memory-based decision-making.

To further characterize the spectral profile of this engagement, we examined ERSP maps from the same hippocampal sources (Figure 4). In both hemispheres, low frequency power, including theta-band (4–8 Hz) activity increased within approximately 200 ms after stimulus onset and remained elevated throughout the post-stimulus interval during both the encoding and retrieval phase. ERSP maps were qualitatively similar across retrieval stimulus types, indicating that hippocampal theta engagement is a general feature of task processing at the level of condition-averaged spectral power.

**FIGURE 4.**
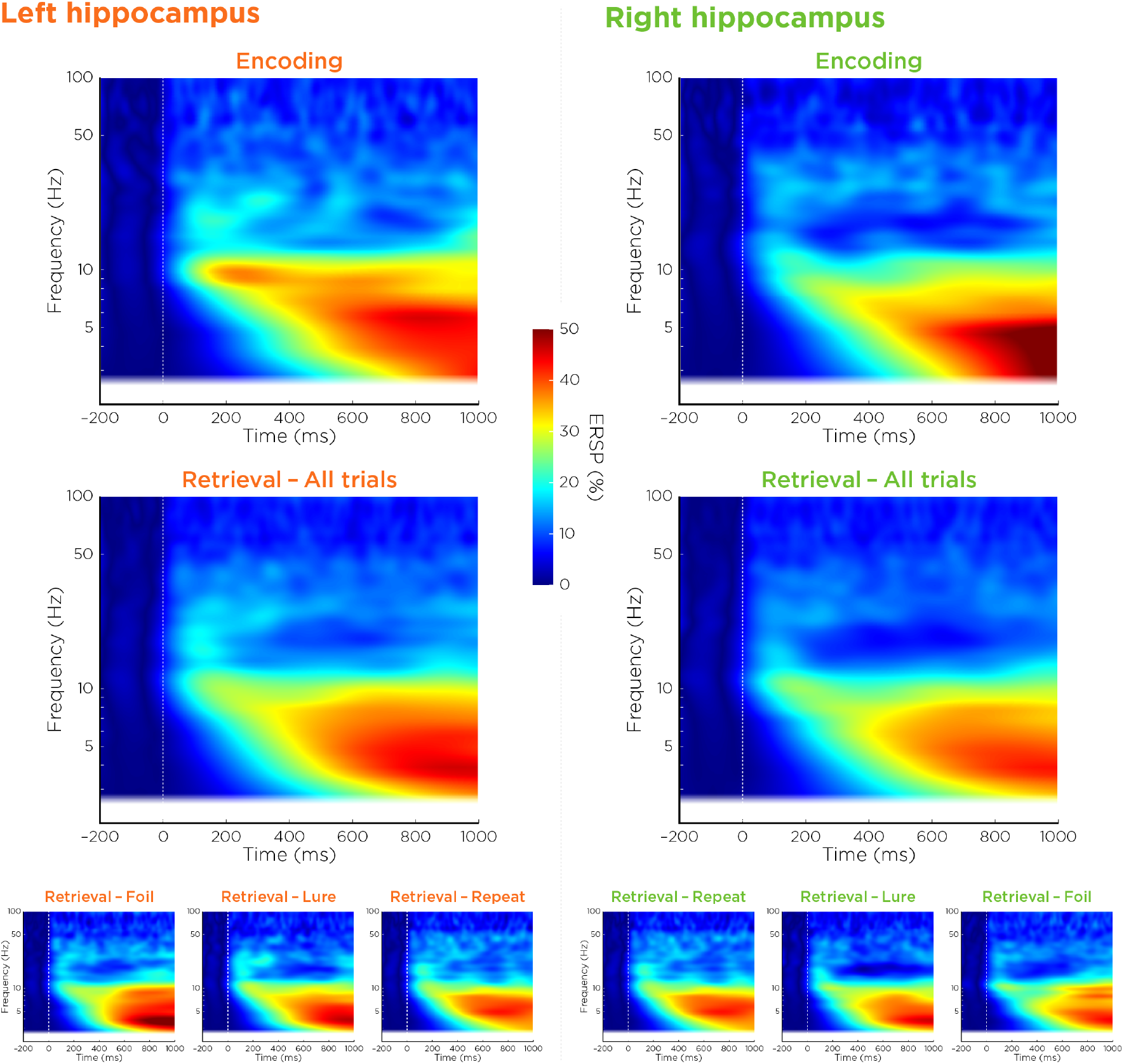
Event-related spectral perturbations (ERSPs) in the left and right hippocampus during encoding and retrieval phases. Time–frequency representations of source-localized hippocampal activity are shown separately for the left (orange labels) and right (green labels) hippocampus. Top row: ERSPs during the encoding phase. Middle row: ERSPs during the retrieval phase, averaged across all trials. Bottom row: Retrieval-phase ERSPs separated by stimulus condition (repeat, lure, foil). Power is expressed as percent change relative to a 200-ms pre-stimulus baseline. Across both task phases and hemispheres, stimuli elicited a sustained increase in low-frequency power, with prominent theta-band (4–8 Hz) engagement emerging within approximately 200 ms after stimulus onset and remaining elevated through the end of the analysis window (up to 1000 ms).

Consistent with these time–frequency patterns, mean theta power was elevated relative to baseline in nearly all stimulus–response conditions (Figure 5), with the strongest increases observed in the left hippocampus for “Old” responses. Bootstrap confidence intervals confirmed that these effects were robust across participants. These descriptive analyses provide a foundation for subsequent single-trial modeling of theta-related decision dynamics.

**FIGURE 5.**
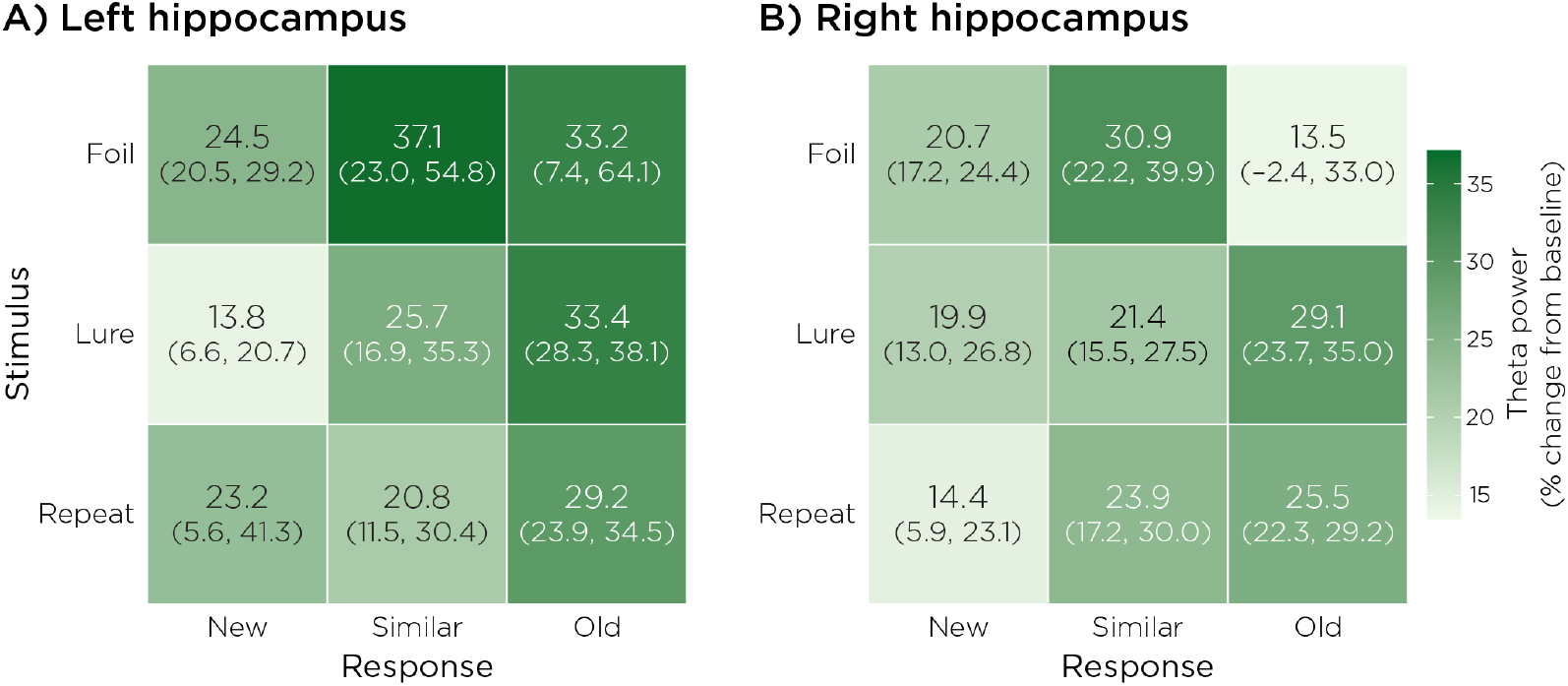
Mean theta power (4–8 Hz) in the hippocampus across stimulus–response pairings. Each cell represents the group-averaged theta power (expressed as percent change from 200 ms pre-stimulus baseline). Parentheses denote 95% bootstrap confidence intervals across participants. Theta power was elevated above baseline in nearly all conditions, indicating that hippocampal theta engagement is a general feature of retrieval-phase processing during the Mnemonic Similarity Task.

### 3.3 Theta-Dependent Drift Rates

We examined whether trial-by-trial hippocampal theta power modulated evidence accumulation (Figure 6). Most theta effects were small: 16 of 18 beta coefficients had 95% credible intervals overlapping zero. However, two effects were credibly nonzero. Left hippocampal theta was associated with a reduction in drift toward “New” responses on Lure trials (*β* = –0.049, 95% CrI [–0.098, –0.0014]). In contrast, right hippocampal theta was associated with an increase in drift toward “Similar” responses on Foil trials (*β* = 0.056, 95% CrI [0.013, 0.098]).

**FIGURE 6.**
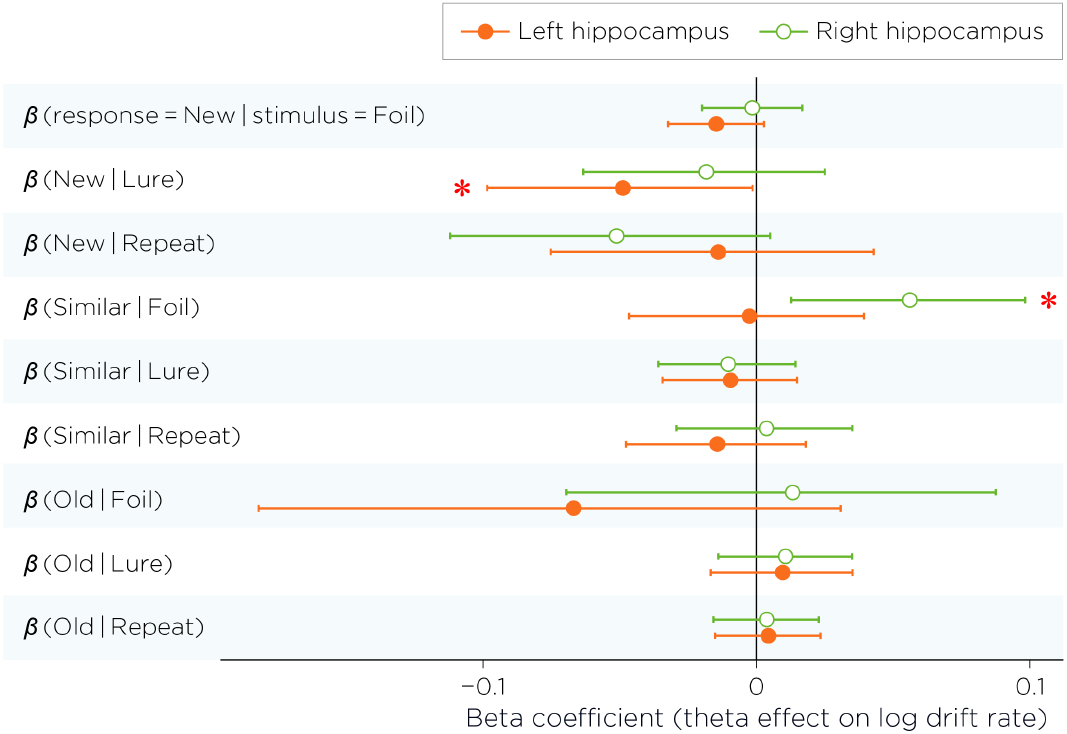
Posterior mean beta coefficients (*β*) representing the effect of trial-by-trial hippocampal theta power on log drift rates for each response–stimulus combination, shown separately for left (orange filled circles) and right (green open circles) hippocampus. Error bars represent 95% credible intervals. Since theta power was *z*-scored, coefficients represent the change in log drift rate per standard deviation increase in theta power. Positive values indicate that higher theta power increases evidence accumulation for that response option. Asterisks indicate effects whose 95% credible intervals exclude zero: left hippocampal theta significantly reduced evidence for “New” responses to Lures (*β* = –0.049, 95% CrI [–0.098, –0.0014]), while right hippocampal theta significantly increased evidence for “Similar” responses to Foils (*β* = 0.056, 95% CrI [0.013, 0.098]). Most theta effects were centered near zero, suggesting limited modulation of drift rates by hippocampal theta oscillation in this task. The vertical line at zero represents the null hypothesis of no theta effect.

Formal tests of hemispheric lateralization (Δ*β* = *β*_left_ – *β*_right_) yielded no credible differences for any response–stimulus combination. Thus, hippocampal theta showed selective, error-linked modulation of evidence accumulation, but no evidence that these theta–drift relationships were systematically lateralized.

### 3.4 LBA Model Evaluation

Posterior sampling converged well for all parameters (max 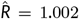, minimum effective sample size = 3, 546, 0 divergent transitions), and trace plots indicated adequate mixing across all four chains (Supplementary Figure S1; Supplementary Table S1). PSIS-LOO cross-validation suggested good out-of-sample predictive accuracy (ELPD = –3498.4, SE = 82.8; Supplementary Section S2). Pareto-*k* diagnostics indicated reliable importance sampling for nearly all observations: 4,905 of 4,907 trials (99.96%) had *k* < 0.7 (Supplementary Figure S2). The remaining two trials exceeded this threshold and were handled using exact LOO refitting (reloo) / moment matching; these trials did not materially change the ELPD estimate (Supplementary Section S2).

Posterior predictive checks indicated that the model adequately reproduced both the RT distributions and choice proportions across stimulus–response combinations, with minor discrepancies for infrequent response types such as “Old” responses to Foils (see Supplementary Figure S3).

### 3.5 Theta Power and Response Times

To evaluate whether theta power was associated with behavioral response speed independently of latent decision processes, we conducted a linear mixed-effects analysis of RTs. We modeled RT as a function of theta power, response type, and stimulus type, including all two- and three-way interactions.

RTs varied significantly across response and stimulus conditions (response: *t* = 5.14, *p* < .001; stimulus: *t* = 5.46, *p* < .001; response *×* stimulus: *t* = –5.76, *p* < .001). However, no main effect or interaction involving theta power reached significance (all *p* > .25; main effect: *t* = –0.24, *p* = .812; theta *×* response: *t* = 0.19, *p* = .846; theta *×* stimulus: *t* = 1.13, *p* = .260; three-way interaction: *t* = –0.90, *p* = .369).

These null results suggest that theta power influences internal decision dynamics that are not detectable in mean RT alone, underscoring the added value of drift-rate modeling in uncovering neurocognitive mechanisms of behavior.

### 3.6 SME Analysis

SME analyses did not reveal significant differences in encoding theta power between later-remembered (Hits) and later-forgotten (Misses) items. Paired *t*-tests across 14 participants did not reveal significant SME effects in either hemisphere (Left: *t* = –0.54, *p* = .601; Right: *t* = 1.28, *p* = .222).

## 4 DISCUSSION

The present study builds on recent efforts to model memory-based decisions using formal evidence accumulation frameworks. In particular, Banavar et al. (Banavar et al., 2024) applied the LBA model to the MST, demonstrating that drift rate—a parameter indexing the quality and speed of evidence accumulation—offers a mechanistically interpretable alternative to surface-level behavioral metrics such as RT (see Section 3.5). Our estimated drift rates exhibited a clear stimulus-congruency structure: the largest drifts occurred for the response option that matched the stimulus class (Figure 2, diagonal; “Old”|Repeat, “Similar”|Lure, “New”|Foil), while off-diagonal accumulators showed markedly weaker evidence accumulation (“Old”|Foil, “New”|Repeat). This pattern supports the interpretation that drift rate tracks the strength of mnemonic evidence driving each decision. Notably, lure trials occupied an intermediate regime, consistent with greater ambiguity and competition among response alternatives. Building on this model-based account, we next asked whether trial-to-trial fluctuations in hippocampal theta power predict the dynamics of evidence accumulation during individual mnemonic decisions.

While hippocampal theta power was broadly elevated across nearly all stimulus–response conditions during the retrieval phase of the MST, this global increase alone does not account for the observed variation in behavioral performance. Average-level effects, though indicative of general retrieval-related engagement, obscure the functional specificity of theta activity in shaping decisions. Notably, our ERSP analyses revealed no condition-specific differences across the full frequency spectrum examined (1–100 Hz), suggesting that mnemonic specificity does not emerge from average oscillatory profiles. This pattern parallels prior MEG studies: Riggs et al. used Synthetic Aperture Magnetometry to examine theta power during complex scene recognition and found no significant differences between old and new stimuli, suggesting that hippocampal theta may reflect a general recognition-related process rather than selective responses to novelty or familiarity (Riggs et al., 2009), though our task affords finer mnemonic distinctions across three stimulus classes. Together, these null average-level findings motivated a shift from condition-averaged to trial-level analyses, where individual fluctuations in theta power could be linked directly to accumulation dynamics. This approach revealed that, even against the backdrop of globally elevated theta, only a subset of stimulus–response pairings showed reliable theta–drift relationships, indicating that it is not merely the presence of theta, but its trial-to-trial fluctuations, that track the dynamic cognitive processes underlying memory-based decisions. This predictive specificity aligns with recent physiological evidence that theta in the human hippocampus occurs in sparse, intermittent bursts rather than as continuous oscillations, even during memory tasks (Penner et al., 2022), reinforcing the importance of trial-level analyses in uncovering the functional role of theta dynamics.

A central finding of the present study was that trial-by-trial hippocampal theta power was associated with *selective* changes in evidence accumulation during mnemonic decisions. Of the 18 theta–drift coefficients estimated across stimulus–response pairings and hemispheres, only two were credibly nonzero. Theta power, therefore, did not exert a broad, uniform influence on evidence accumulation. Rather, its effects emerged in specific contexts under specific stimulus demands. Notably, both credible effects involved partial match judgments, but the direction of modulation differed across contexts. Higher left hippocampal theta was associated with *reduced* drift toward “New” responses on Lure trials. For these trials, the stimulus shares partially overlapping features with a previously encoded item, where a “New” response represents a discrimination failure. Elevated theta was associated with reduced drift toward an erroneous novelty judgment, consistent with sensitivity to partial mnemonic overlap. Although drift rate aggregates multiple cognitive influences and we cannot rule out contributions from attentional or criterion shifts, the selectivity of this effect to Lure trials—where partial overlap is the defining feature—favors a mnemonic interpretation. Conversely, higher right hippocampal theta was associated with increased drift toward “Similar” responses on Foil trials, where no encoded memory counterpart exists. This suggests that the same partial-match sensitivity can amplify spurious similarity signals when no true memory match is present. An alternative account in terms of general conflict resolution cannot be excluded, though this would not obviously predict the specificity of the effect to Foil trials alone. Together, these findings suggest that hippocampal theta is selectively associated with evidence accumulation in contexts involving partial stimulus overlap. Whether this association is beneficial or costly depends on the stimulus context: when overlap reflects a genuine memory, the theta–drift relationship favors correct discrimination; when overlap is absent, the same relationship is linked to an erroneous response, suggesting that the behavioral consequences of hippocampal theta fluctuations are shaped by the mnemonic quality of the evidence present on a given trial.

Although the two credible effects arose in different hemispheres, direct contrasts between left and right coefficients were not credibly different from zero for any stimulus–response pairing. We therefore treat the hemispheric pattern as descriptive rather than as evidence for functional asymmetry. This cautious stance nonetheless leaves open the possibility of hemisphere-specific contributions, given intracranial findings that hippocampal theta properties differ between hemispheres (Penner et al., 2022) and neuroimaging work suggesting hemispheric and subfield gradients in mnemonic discrimination (Stevenson et al., 2020). Whether hemisphere-specific theta dynamics contribute meaningfully to these effects remains an open question, one that future work incorporating subfield-resolved recordings and larger samples would be better positioned to address.

We note that encoding-phase hippocampal theta did not significantly predict subsequent memory in either hemisphere. This null SME is consistent with the mnemonic similarity task design, which emphasizes fine-grained retrieval discrimination rather than overall encoding strength, and may reflect that encoding effects are subfield-specific (Bakker et al., 2008) and, thus, undetectable with whole-hippocampus source localization. The extended task design, as noted above, may have also contributed to this null result, with which the prolonged retrieval period could introduce greater interference than typical SME paradigms. Critically, this dissociation between encoding and retrieval theta underscores that our retrieval findings reflect a mechanistically distinct process wherein theta dynamically guides evidence accumulation during decision-making rather than simply indexing memory strength.

Although hippocampal sources are challenging to resolve with MEG, our ERFs analysis demonstrates that temporally reliable, stimulus-locked activity can be detected from this deep brain structure. Group-averaged RMS time courses revealed a consistent response pattern in both hippocampi during both the encoding and retrieval phase, characterized by an initial deflection beginning around 100 ms after stimulus onset, a polyphasic wave peaking between 200–300 ms, and a smaller subsequent peak between 400–500 ms. This temporal profile closely aligns with previous MEG findings of hippocampal activity during recognition memory tasks (Anderson et al., 2017, Gonsalves et al., 2005, Riggs et al., 2009). The early onset of this response suggests that hippocampal contributions to recognition begin rapidly, potentially in parallel with visual processing (Riggs et al., 2009), while the later sustained component likely reflects ongoing mnemonic evaluation processes such as recollection (Gonsalves et al., 2005, Riggs et al., 2009). These results add to growing evidence that MEG, when combined with appropriate source modeling and localization methods, can reliably detect signals from deep brain structures such as the hippocampus (Bénar et al., 2021, López-Madrona et al., 2022, Pizzo et al., 2019). Although our RMS-based aggregation across dipoles precludes subfield-level analysis, it provides a robust estimate of hippocampal engagement suitable for assessing condition-related evoked dynamics.

While our findings underscore the utility of combining computational modeling with source-localized MEG, several methodological considerations warrant caution. The mean recognition score in the present study (0.57) was lower than values reported in prior MST studies (typically 0.7–0.8 (Stark et al., 2019)). This reduction likely reflects our extended retrieval phase (360 trials vs. 192 in standard protocols), which was necessary to achieve sufficient trial counts for reliable MEG source localization but increased proactive interference and cognitive demands. Consistent with this interpretation, repeat hit rates (64%) were lower than typical benchmarks, while foil correct rejection rates (93%) remained high (Figure 2A), suggesting that reduced recognition primarily reflected memory decay and interference rather than general task confusion. Additional contributors may include differences in experimental context. Notably, the task was administered during MEG recording, which involves head stabilization, projector presentation of the stimulus, and longer overall session duration; these factors may modestly affect attention, response caution, and perceptual confidence. Importantly, stimulus-type effects remained robust and significant, indicating that despite lower absolute performance, the task reliably engaged mnemonic discrimination processes.

Although our sample size (*N* = 14) was sufficient to support robust within-subject modeling of neural and behavioral dynamics, the demographics may limit generalizability. The sample was predominantly female (13 of 14 participants), which may introduce bias given known sex differences in episodic memory and pattern separation (Herlitz et al., 1997, Yagi et al., 2016). Additionally, the cohort was relatively young (mean age 32), and age-related changes in hippocampal circuitry and pattern separation performance (Riphagen et al., 2020, Stark et al., 2013, Yassa & Stark, 2011) may constrain the applicability of these findings to older populations. Further, although recent work has validated the feasibility of localizing hippocampal sources with MEG, such estimates remain constrained by signal-to-noise ratios, inverse modeling assumptions, and anatomical variability. We employed conservative localization procedures based on individualized MRI models and constrained dSPM estimates. Our interpretations remain at the level of whole hippocampal dynamics. This whole-hippocampus approach was necessitated by the spatial resolution constraints of MEG for deep sources: beamformer localization of hippocampal signals shows errors of 7–15 mm even under optimal conditions (Quraan et al., 2011), approaching the spatial extent of anterior–posterior subdivisions and precluding confident rostrocaudal differentiation. Emerging techniques, such as the reciprocal boundary element fast multipole method (BEM-FMM) (Drumm et al., 2025, Wartman et al., 2025), offer promising avenues for improving spatial specificity in future studies.

While the LBA model offers a principled decomposition of decision processes, it necessarily abstracts over factors such as strategic shifts or attentional variability. Additionally, while the 0–1000 ms post-stimulus window was selected based on prior work and the observed RT distribution, we cannot fully exclude contributions from perceptual or incidental encoding activity within this interval; response-locked analyses or designs with trial-level encoding–retrieval separation would be needed to more precisely isolate decision-related theta activity. Nevertheless, by linking trial-level neural activity to model-derived indices of evidence accumulation, our approach advances the integration of cognitive theory with temporally precise neurophysiological data.

Looking forward, this integrative framework opens new avenues for investigating the neural dynamics of memory-guided decision-making. Future work could extend these findings to populations with known impairments in hippocampal function, such as older adults, individuals with mild cognitive impairment, or those with affective disorders, in whom pattern separation deficits are well documented (Déry et al., 2013, Stark et al., 2013). Moreover, causal interventions targeting theta oscillations, for example, via transcranial brain stimulation Deng et al. (2015), pharmacological modulation, or behavioral entrainment, could further elucidate the functional role of theta in biasing hippocampal computations. Finally, expanding this approach to other neural markers or brain regions may refine our understanding of how distributed circuits coordinate under conditions of uncertainty. By combining fine-grained behavioral modeling with temporally resolved neurophysiology, we move closer to a mechanistic account of how memory is transformed into a decision.

## 5 CONCLUSIONS

Hippocampal theta oscillations appear to selectively track mnemonic evidence accumulation in contexts involving partial memory overlap, though their influence across the broader range of stimulus–response pairings was limited. These findings suggest that theta does not uniformly drive memory-guided decisions, but may instead play a targeted role in regulating sensitivity to mnemonic similarity. By combining non-invasive deep-source MEG with hierarchical computational modeling, this work offers a methodological framework for bridging neural oscillatory dynamics and latent decision processes. Future studies with larger and more diverse samples, and with causal tools to manipulate theta directly, will be needed to clarify the functional significance and hemispheric organization of these effects, particularly in populations where hippocampal integrity is compromised.

## Data Availability

All data produced in the present study are available upon reasonable request to the authors.

## AUTHOR CONTRIBUTIONS

Conceptualization, Z.-D.D. and P.L.R.; methodology, P.L.R., J.R.G., and Z.-D.D.; validation, P.L.R., J.D.S., F.W.C., and Z.-D.D.; formal analysis, P.L.R. and Z.-D.D.; data curation, J.R.G., N.M., P.L.R., and Z.-D.D.; writing–original draft preparation, P.L.R. and Z.-D.D.; writing–review and editing, P.L.R., J.R.G., B.L., N.M., E.B., J.D.S., F.W.C., and Z.-D.D.; visualization, P.L.R. and Z.-D.D.; supervision, Z.-D.D. All authors have read and agreed to the published version of the manuscript.

## ACKNOWLEDGMENTS

This research was supported by the Intramural Research Program of the National Institutes of Health (NIH). The contributions of the NIH authors were made as part of their official duties as NIH federal employees, are in compliance with agency policy requirements, and are considered Works of the United States Government. However, the findings and conclusions presented in this paper are those of the authors and do not necessarily reflect the views of the NIH or the U.S. Department of Health and Human Services.

Computational analyses were performed using the NIH HPC Biowulf cluster (http://hpc.nih.gov). The authors thank the NIMH MEG Core Facility for their contributions to data acquisition and technical support. We thank Alexandra Halberstadt for programming the mnemonic task in PsychoPy.

## REFERENCES

Addante, R. J., Watrous, A. J., Yonelinas, A. P., Ekstrom, A. D., & Ranganath, C. (2011). Prestimulus theta activity predicts correct source memory retrieval. Proc Natl Acad Sci U S A, 108(26), 10702–10707. doi: 10.1073/pnas.1014528108

Allegra, M., Posani, L., Gómez-Ocádiz, R., & Schmidt-Hieber, C. (2020). Differential relation between neuronal and behavioral discrimination during hippocampal memory encoding. Neuron, 108(6), 1103–1112. doi: 10.1016/j.neuron.2020.09.032

Anderson, M. L., James, J. R., & Kirwan, C. B. (2017). An event-related potential investigation of pattern separation and pattern completion processes. Cogn Neurosci, 8(1), 9–23. doi: 10.1080/17588928.2016.1195804

Attal, Y., Bhattacharjee, M., Yelnik, J., Cottereau, B., Lefèvre, J., Okada, Y., … Baillet, S. (2009). Modelling and detecting deep brain activity with MEG and EEG. IRBM, 30(3), 133–138. doi: 10.1016/j.irbm.2009.01.005

Attal, Y., & Schwartz, D. (2013). Assessment of subcortical source localization using deep brain activity imaging model with minimum norm operators: A MEG study. PLoS One, 8(3), e59856. doi: 10.1371/journal.pone.0059856

Bakker, A., Kirwan, C. B., Miller, M., & Stark, C. E. L. (2008). Pattern separation in the human hippocampal CA3 and dentate gyrus. Science, 319(5870), 1640–1642. doi: 10.1126/science.1152882

Banavar, N. V., Noh, S. M., Wahlheim, C. N., Cassidy, B. S., Kirwan, C. B., Stark, C. E. L., & Bornstein, A. M. (2024). A response time model of the three-choice Mnemonic Similarity Task provides stable, mechanistically interpretable individual-difference measures. Front Hum Neurosci, 18, 1379287. doi: 10.3389/fnhum.2024.1379287

Bénar, C.-G., Velmurugan, J., López-Madrona, V. J., Pizzo, F., & Badier, J.-M. (2021). Detection and localization of deep sources in magnetoencephalography: A review. Curr Opin Biomed Eng, 18, 100285. doi: 10.1016/j.cobme.2021.100285

Berron, D., Schütze, H., Maass, A., Cardenas-Blanco, A., Kuijf, H. J., Kumaran, D., & Düzel, E. (2016). Strong evidence for pattern separation in human dentate gyrus. Journal of Neuroscience, 36(29), 7569–7579. doi: 10.1523/JNEUROSCI.0518-16.2016

Betancourt, M., & Girolami, M. (2015). Hamiltonian monte carlo for hierarchical models. In S. K. Upadhyay, U. Singh, D. K. Dey, & A. Loganathan (Eds.), Current trends in bayesian methodology with applications. New York: Chapman and Hall/CRC.

Brown, S. D., & Heathcote, A. (2008). The simplest complete model of choice response time: Linear ballistic accumulation. Cogn Psychol, 57(3), 153–178. doi: 10.1016/j.cogpsych.2007.12.002

Buzsáki, G. (2002). Theta oscillations in the hippocampus. Neuron, 33(3), 325–340. doi: 10.1016/s0896-6273(02)00586-x

Colgin, L. L. (2013). Mechanisms and functions of theta rhythms. Annu Rev Neurosci, 36, 295–312. doi: 10.1146/annurev-neuro-062012-170330

Dale, A. M., Liu, A. K., Fischl, B. R., Buckner, R. L., Belliveau, J. W., Lewine, J. D., & Halgren, E. (2000). Dynamic statistical parametric mapping: combining fMRI and MEG for high-resolution imaging of cortical activity. Neuron, 26(1), 55–67. doi: 10.1016/s0896-6273(00)81138-1

Deng, Z.-D., McClintock, S. M., Oey, N. E., Luber, B., & Lisanby, S. H. (2015). Neuromodulation for mood and memory: from the engineering bench to the patient bedside. Curr Opin Neurobiol, 30, 38–43. doi: 10.1016/j.conb.2014.08.015

Déry, N., Pilgrim, M., Gibala, M., Gillen, J., Wojtowicz, J. M., MacQueen, G., & Becker, S. (2013). Adult hippocampal neurogenesis reduces memory interference in humans: Opposing effects of aerobic exercise and depression. Front Neurosci, 7, 66. doi: 10.3389/fnins.2013.00066

Donkin, C., Brown, S., & Heathcote, A. (2011). Drawing conclusions from choice response time models: A tutorial using the linear ballistic accumulator. J Math Psychol, 55(2), 140–151. doi: 10.1016/j.jmp.2010.10.001

Donkin, C., Brown, S., Heathcote, A., & Wagenmakers, E.-J. (2011). Diffusion versus linear ballistic accumulation: Different models but the same conclusions about psychological processes? Psychon Bull Rev, 18(1), 61–69. doi: 10.3758/s13423-010-0022-4

Drumm, D. A., Nuñez Ponasso, G., Linke, A., Noetscher, G. M., Maess, B., Knösche, T. R., … Deng, Z.-D. (2025). Improved source localization of auditory evoked fields using reciprocal BEM-FMM. bioRxiv. doi: 10.1101/2025.05.09.653081

Eichenbaum, H. (2017). Prefrontal–hippocampal interactions in episodic memory. Nat Rev Neurosci, 18(9), 547–558. doi: 10.1038/nrn.2017.74

Engel, A. K., & Fries, P. (2010). Beta-band oscillations — signalling the status quo? Curr Opin Neurobiol, 20(2), 156–165. doi: 10.1016/j.conb.2010.02.015

Fell, J., Ludowig, E., Staresina, B. P., Wagner, T., Kranz, T., Elger, C. E., & Axmacher, N. (2011). Medial temporal theta/alpha power enhancement precedes successful memory encoding: Evidence from intracranial EEG. J Neurosci, 31(14), 5392–5397. doi: 10.1523/JNEUROSCI.3668-10.2011

Fischl, B. (2013). FreeSurfer. Neuroimage, 62(2), 774–781. doi: 10.1016/j.neuroimage.2012.01.021

Gelman, A., & Rubin, D. B. (1992). Inference from iterative simulation using multiple sequences. Statist Sci, 7(4), 457–472. doi: 10.1214/ss/1177011136

Gonsalves, B. D., Kahn, I., Curran, T., Norman, K. A., & Wagner, A. D. (2005). Memory strength and repetition suppression: Multimodal imaging of medial temporal cortical contributions to recognition. Neuron, 47(5), 751–761. doi: 10.1016/j.neuron.2005.07.013

Guderian, S., Schott, B. H., Richardson-Klavehn, A., & Düzel. (2009). Medial temporal theta state before an event predictsepisodic encoding success in humans. Proc Natl Acad Sci U S A, 106(13), 5365–5370. doi: 10.1073/pnas.0900289106

Hasselmo, M. E. (2005). What is the function of hippocampal theta rhythm?–Linking behavioral data to phasic properties of field potential and unit recording data. Hippocampus, 15(7), 936–949. doi: 10.1002/hipo.20116

Hasselmo, M. E., Bodelón, C., & Wyble, B. P. (2002). A proposed function for hippocampal theta rhythm: Separate phases of encoding and retrieval enhance reversal of prior learning. Neural Comput, 14(4), 793–817. doi: 10.1162/089976602317318965

Herlitz, A., Nilsson, L.-G., & Bäckman, L. (1997). Gender differences in episodic memory. Mem Cognit, 25(6), 801–811. doi: 10.3758/bf03211324

Herweg, N. A., Solomon, E. A., & Kahana, M. J. (2020). Theta oscillations in human memory. Trends Cogn Sci, 24(3), 208–227. doi: 10.1016/j.tics.2019.12.006

Klimesch, W. (2012). Alpha-band oscillations, attention, and controlled access to stored information. Trends Cogn Sci, 16(12), 602–617. doi: 10.1016/j.tics.2012.10.007

Leutgeb, J. K., Leutgeb, S., Moser, M.-B., & Moser, E. I. (2007). Pattern separation in the dentate gyrus and CA3 of the hippocampus. Science, 315(5814), 961–966. doi: 10.1126/science.1135801

López-Madrona, V. J., Medina Villalon, S., Badier, J. M., Tréuchon, A., Jayabal, V., Bartolomei, F., … Bénar, C. G. (2022). Magnetoencephalography can reveal deep brain network activities linked to memory processes. Hum Brain Mapp, 43(15), 4733–4749. doi: 10.1002/hbm.25987

Mueller, C. J., White, C. N., & Kuchinke, L. (2017). Electrophysiological correlates of the drift diffusion model in visual word recognition. Hum Brain Mapp, 38(11), 5616–5627. doi: 10.1002/hbm.23753

Mulder, M. J., van Maanen, L., & Forstmann, B. U. (2014). Perceptual decision neurosciences - a model-based review. Neuroscience, 277, 872–884. doi: 10.1016/j.neuroscience.2014.07.031

O’Reilly, R. C., & McClelland, J. L. (1994). Hippocampal conjunctive encoding, storage, and recall: Avoiding a trade-off. Hippocampus, 4(5), 661–682. doi: 10.1002/hipo.450040605

Osipova, D., Takashima, A., Oostenveld, R., Fernández, G., Maris, E., & Jensen, O. (2006). Theta and gamma oscillations predict encoding and retrieval of declarative memory. J Neurosci, 26(28), 7523–7531. doi: 10.1523/JNEUROSCI.1948-06.2006

Pacheco Estefan, D., Zucca, R., Arsiwalla, X., Principe, A., Zhang, H., Rocamora, R., … Verschure, P. F. M. J. (2021). Volitional learning promotes theta phase coding in the human hippocampus. Proc Natl Acad Sci U S A, 118(10), e2021238118. doi: 10.1073/pnas.2021238118

Peirce, J., Gray, J. R., Simpson, S., MacAskill, M., Höchenberger, R., Sogo, H., … Lindeløv, J. K. (2019). PsychoPy2: Experiments in behavior made easy. Behav Res Methods, 51(1), 195–203. doi: 10.3758/s13428-018-01193-y

Penner, C., Minxha, J., Chandravadia, N., Mamelak, A. N., & Rutishauser, U. (2022). Properties and hemispheric differences of theta oscillations inthe human hippocampus. Hippocampus, 32(5), 335–341. doi: 10.1002/hipo.23412

Philiastides, M. G., & Sajda, P. (2006). Temporal characterization of the neural correlates of perceptual decision making in the human brain. Cereb Cortex, 16(4), 509–518. doi: 10.1093/cercor/bhi130

Pizzo, F., Roehri, N., Medina Villalon, S., Tréuchon, A., Chen, S., Lagarde, S., … Bénar, C. G. (2019). Deep brain activities can be detected with magnetoencephalography. Nat Commun, 10, 971. doi: 10.1038/s41467-019-08665-5

Quraan, M. A., Moses, S. N., Hung, Y., Mills, T., & Taylor, M. J. (2011). Detection and localization of hippocampal activity using beamformers with meg: a detailed investigation using simulations and empirical data. Hum Brain Mapp, 32(5), 812–827. doi: 10.1002/hbm.21068

Ratcliff, R., Philiastides, M. G., & Sajda, P. (2009). Quality of evidence for perceptual decision making is indexed by trial-to-trial variability of the EEG. Proc Natl Acad Sci U S A, 106(16), 6539–6544. doi: 10.1073/pnas.0812589106

Riggs, L., Moses, S. N., Bardouille, T., Herdman, A. T., Ross, B., & Ryan, J. D. (2009). A complementary analytic approach to examining medial temporal lobe sourcesusing magnetoencephalography. Neuroimage, 45(2), 627–642. doi: 10.1016/j.neuroimage.2008.11.018

Riphagen, J. M., Schmiedek, L., Gronenschild, E. H. B. M., Yassa, M. A., Priovoulos, N., Sack, A. T., … Jacobs, H. I. L. (2020). Associations between pattern separation and hippocampal subfield structure and function vary along the lifespan: A 7 T imaging study. Sci Rep, 10(1), 7572. doi: 10.1038/s41598-020-64595-z

Rolls, E. T. (2013). The mechanisms for pattern completion and pattern separation in the hippocampus. Front Syst Neurosci, 7, 74. doi: 10.3389/fnsys.2013.00074

Saint Amour di Chanaz, L., Pérez-Bellido, A., Wu, X., Lozano-Soldevilla, D., Pacheco-Estefan, D., Lehongre, K., … Fuentemilla, L. (2023). Gamma amplitude is coupled to opposed hippocampal theta-phase states during the encoding and retrieval of episodic memories in humans. Curr Biol, 33(9), 1836–1843.e6. doi: 10.1016/j.cub.2023.03.073

Stan Development Team. (2024). RStan: The R interface to Stan. Retrieved from https://mc-stan.org/ R package version 2.26.24.

Stark, S. M., Kirwan, C. B., & Stark, C. E. L. (2019). Mnemonic similarity task: A tool for assessing hippocampal integrity. Trends Cogn Sci, 23(11), 938–951. doi: 10.1016/j.tics.2019.08.003

Stark, S. M., Yassa, M. A., Lacy, J. W., & Stark, C. E. L. (2013). A task to assess behavioral pattern separation (BPS) in humans: Data from healthy aging and mild cognitive impairment. Neuropsychologia, 51(12), 2442–2449. doi: 10.1016/j.neuropsychologia.2012.12.014

Stevenson, R. F., Reagh, Z. M., Chun, A. P., Murray, E. A., & Yassa, M. A. (2020). Pattern separation and source memory engage distinct hippocampal and neocortical regions during retrieval. J Neurosci, 40(4), 843–851. doi: 10.1523/JNEUROSCI.0564-19.2019

Tadel, F., Baillet, S., Mosher, J. C., Pantazis, D., & Leahy, R. M. (2011). Brainstorm: A user-friendly application for MEG/EEG analysis. Comput Intell Neurosci, 2011, 879716. doi: 10.1155/2011/879716

Treves, A., & Rolls, E. T. (1994). Computational analysis of the role of the hippocampus in memory. Hippocampus, 4(3), 374–391. doi: 10.1002/hipo.450040319

Tulving, E. (1983). Elements of Episodic Memory. Oxford, UK: Oxford University Press.

Tulving, E., Kapur, S., Craik, F. I., Moscovitch, M., & Houle, S. (1994). Hemispheric encoding/retrieval asymmetry in episodic memory: Positron emission tomography findings. Proc Natl Acad Sci U S A, 91(6), 2016–2020. doi: 10.1073/pnas.91.6.2016

Vehtari, A., Gelman, A., & Gabry, J. (2017). Practical Bayesian model evaluation using leave-one-out cross-validation and WAIC. Stat Comput, 27, 1413–1432. doi: 10.1007/s11222-016-9696-4

Wartman, W. A., Nuñez Ponasso, G., Qi, Z., Haueisen, J., Maess, B., Knösche, T. R., … Makaroff, S. N. (2025). Fast EEG/MEG BEM-based forward problem solution for high-resolution head models. Neuroimage, 306, 120998. doi: 10.1016/j.neuroimage.2024.120998

Yagi, S., Chow, C., Lieblich, S. E., & Galea, L. A. M. (2016). Sex and strategy use matters for pattern separation, adult neurogenesis, and immediate early gene expression in the hippocampus. Hippocampus, 26(1), 87–101. doi: 10.1002/hipo.22493

Yassa, M. A., & Stark, C. E. L. (2011). Pattern separation in the hippocampus. Trends Neurosci, 34(10), 515–525. doi: 10.1016/j.tins.2011.06.006

Yassa, M. A., Stark, S. M., Bakker, A., Albert, M. S., Gallagher, M., & Stark, C. E. L. (2011). High-resolution structural and functional MRI of hippocampal CA3 and dentate gyrus in patients with amnestic mild cognitive impairment. Neuroimage, 51(3), 1242–1252. doi: 10.1016/j.neuroimage.2010.03.040

