## Supplementary Materials for "Selective Modulation of Evidence Accumulation by Hippocampal Theta Oscillations during Mnemonic Decision-Making"

#### S1 MCMC Convergence and Sampler Diagnostics

To evaluate MCMC convergence and sampling quality for the hierarchical LBA model, we inspected trace plots for representative group-level parameters and summarized standard convergence diagnostics across key parameters. Figure S1 shows trace plots for the group-level location parameters governing the starting-point range $\mu_{\log A}$, threshold separation $\mu_{\log(b-A)}$, and non-decision time $\mu_{\log t_0}$, as well as the across-trial drift-rate variability parameter $s_v$. Across all four chains, these traces exhibit stable stationarity and substantial overlap, indicating adequate mixing and convergence.

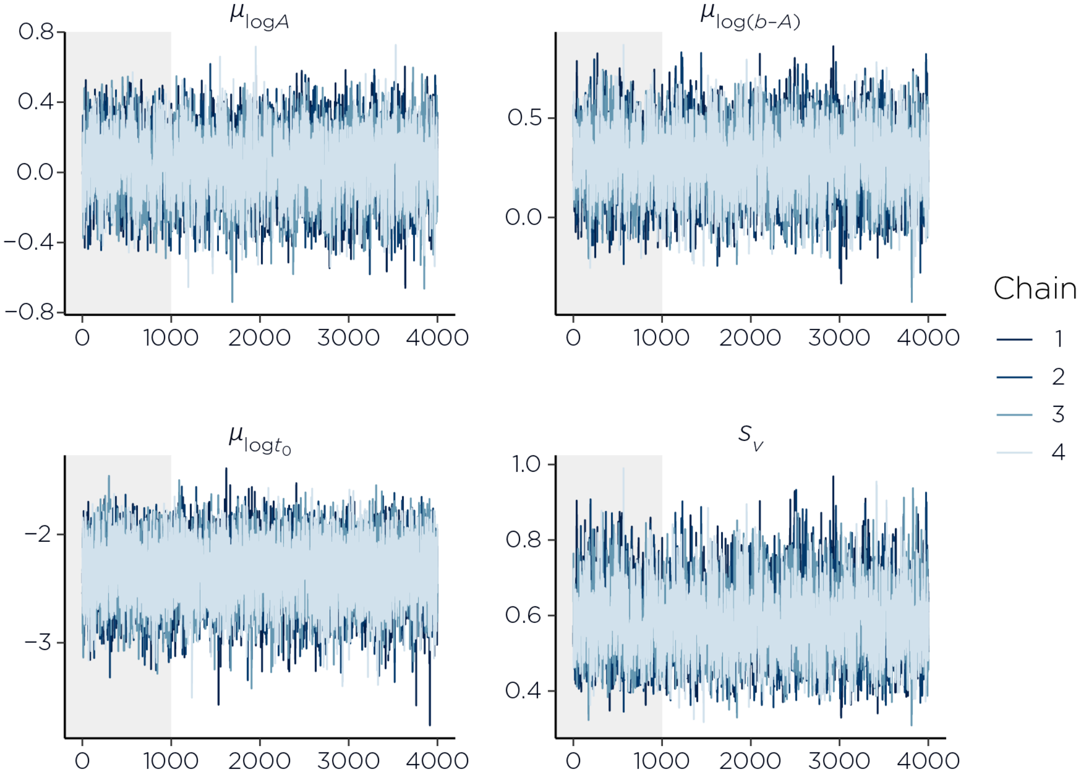


Figure S1: Trace plots demonstrating MCMC convergence for hierarchical model parameters. Trace plots from four independent chains are shown for the group-level parameters $\mu_{\log A}$, $\mu_{\log(b-A)}$, $\mu_{\log t_0}$, and the across-subject variability parameter $(s_v)$. Shaded regions indicate burn-in samples excluded from posterior inference. All chains exhibit good mixing, stationarity, and substantial overlap across chains, with no visible drift or chain-specific behavior, supporting adequate convergence and stable posterior estimation.

Table S1 reports posterior means, posterior standard deviations, central 95% credible intervals, effective sample sizes ($n_{\text{eff}}$), and Gelman–Rubin diagnostics ($\hat{R}$) for key group-level drift-rate parameters $\mu_{\log v[k,s]}$, their corresponding variability terms $\sigma_{\log v[k,s]}$, and the remaining hierarchical parameters $\mu_{\log A}$, $\mu_{\log(b-A)}$, $\mu_{\log t_0}$, $\sigma_{\log A}$, $\sigma_{\log(b-A)}$, $\sigma_{\log t_0}$, and $s_v$. Convergence diagnostics were uniformly satisfactory, with $\hat{R}$ values essentially equal to 1 (maximum $\hat{R} = 1.002$) and large effective sample sizes (minimum $n_{\text{eff}} = 3546$). In addition, Hamiltonian Monte Carlo sampling produced no divergent transitions, supporting stable posterior estimation.

Table S1: Posterior summaries and convergence diagnostics for key group-level and variability parameters in the hierarchical LBA model.

| Parameter | Mean | SD | 2.5% | 97.5% | $n_{\text{eff}}$ | $\hat{R}$ |
|---|---|---|---|---|---|---|
| $\mu_{\log v[1,1]}$ | 0.773 | 0.152 | 0.473 | 1.067 | 3546.008 | 1.000 |
| $\mu_{\log v[1,2]}$ | 0.208 | 0.155 | −0.100 | 0.505 | 3693.833 | 1.000 |
| $\mu_{\log v[1,3]}$ | 0.068 | 0.166 | −0.262 | 0.390 | 3976.680 | 1.001 |
| $\mu_{\log v[2,1]}$ | 0.156 | 0.166 | −0.177 | 0.472 | 4016.968 | 1.000 |
| $\mu_{\log v[2,2]}$ | 0.491 | 0.160 | 0.173 | 0.804 | 3607.776 | 1.001 |
| $\mu_{\log v[2,3]}$ | 0.371 | 0.161 | 0.050 | 0.685 | 3850.248 | 1.000 |
| $\mu_{\log v[3,1]}$ | −0.227 | 0.188 | −0.607 | 0.131 | 5077.759 | 1.000 |
| $\mu_{\log v[3,2]}$ | 0.474 | 0.154 | 0.165 | 0.770 | 3658.883 | 1.001 |
| $\mu_{\log v[3,3]}$ | 0.703 | 0.150 | 0.407 | 0.993 | 3548.059 | 1.001 |
| $\mu_{\log A}$ | 0.048 | 0.175 | −0.308 | 0.379 | 4188.736 | 1.000 |
| $\mu_{\log(b-A)}$ | 0.290 | 0.160 | −0.033 | 0.597 | 4010.603 | 1.001 |
| $\mu_{\log t_0}$ | −2.335 | 0.268 | −2.896 | −1.848 | 10327.963 | 1.000 |
| $\sigma_{\log v[1,1]}$ | 0.131 | 0.035 | 0.077 | 0.212 | 6822.444 | 1.000 |
| $\sigma_{\log v[1,2]}$ | 0.173 | 0.053 | 0.088 | 0.296 | 6401.423 | 1.000 |
| $\sigma_{\log v[1,3]}$ | 0.276 | 0.078 | 0.151 | 0.457 | 6767.516 | 1.001 |
| $\sigma_{\log v[2,1]}$ | 0.290 | 0.078 | 0.168 | 0.472 | 6883.597 | 1.000 |
| $\sigma_{\log v[2,2]}$ | 0.253 | 0.064 | 0.153 | 0.400 | 6328.270 | 1.000 |
| $\sigma_{\log v[2,3]}$ | 0.251 | 0.067 | 0.147 | 0.408 | 6040.329 | 1.000 |
| $\sigma_{\log v[3,1]}$ | 0.384 | 0.105 | 0.216 | 0.621 | 9772.926 | 1.000 |
| $\sigma_{\log v[3,2]}$ | 0.169 | 0.044 | 0.101 | 0.273 | 6409.754 | 1.000 |
| $\sigma_{\log v[3,3]}$ | 0.119 | 0.034 | 0.064 | 0.199 | 5480.521 | 1.000 |
| $\sigma_{\log A}$ | 0.232 | 0.086 | 0.083 | 0.422 | 6045.295 | 1.000 |
| $\sigma_{\log(b-A)}$ | 0.171 | 0.053 | 0.088 | 0.295 | 6894.895 | 1.000 |
| $\sigma_{\log t_0}$ | 0.652 | 0.159 | 0.364 | 0.990 | 9776.133 | 1.000 |
| $s_v$ | 0.580 | 0.089 | 0.422 | 0.770 | 3642.281 | 1.001 |

### S2 LOO Cross-Validation and Influence Diagnostics

We assessed model reliability and influence using PSIS-LOO at both the trial and participant levels. At the trial level, Pareto shape parameters ($k$) indicated stable importance sampling for nearly all observations (Figure S2): only two of 4,907 trials exceeded the conventional $k > 0.7$ threshold. We addressed these trials using exact refitting (`reloo`) / moment matching, and the resulting ELPD estimate did not change materially.

To evaluate whether predictive fit was disproportionately driven by individual participants, we aggregated pointwise PSIS-LOO quantities by summing $\text{elpd}_{\text{loo}}$ and $p_{\text{loo}}$ across each participant's

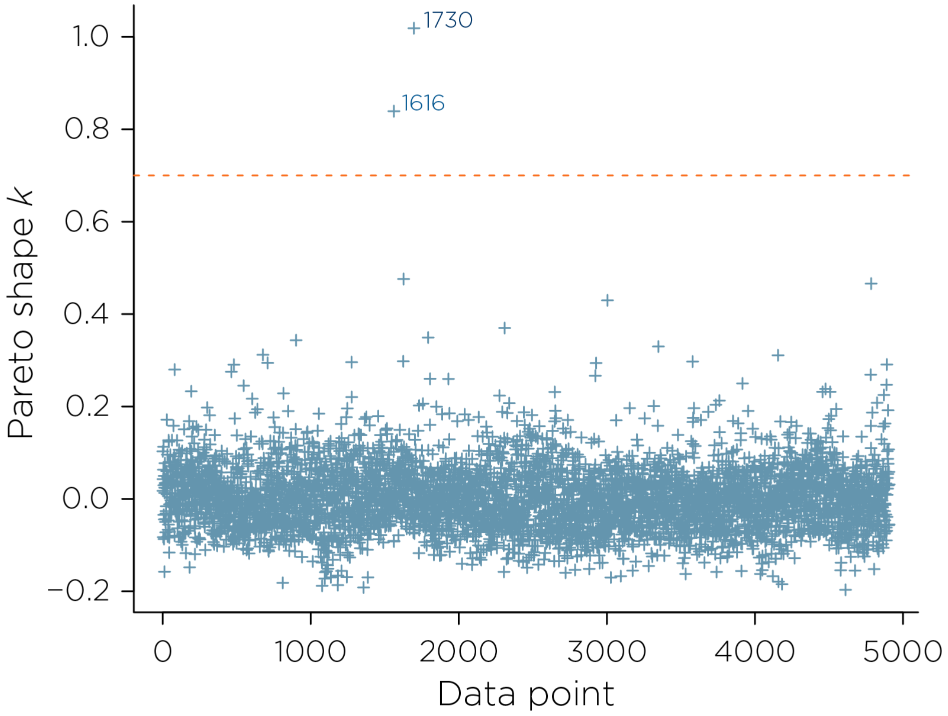


Figure S2: Trial-level Pareto shape parameters ($k$) from leave-one-out cross-validation are shown for each of the 4,907 trials. The dashed line indicates the recommended reliability threshold ($k = 0.7$). The vast majority of observations (4,905; 99.96%) fall below this cutoff, supporting stable importance sampling weights and reliable LOO estimates. Two trials exceeded the threshold; however, their minimal proportion and lack of impact on the overall ELPD indicate negligible influence on model evaluation.

trials (Table S2). Variation in $\text{elpd}_{\text{loo}}$ across participants (range: $-387$ to $-145$) is consistent consistent with expected individual differences in response patterns and trial count. One participant (Participant 5) showed a notably lower $\text{elpd}_{\text{loo}}$ ($-387$) and elevated $p_{\text{loo}}$ (23.4) relative to the remaining participants, suggesting their data required greater model complexity to fit. However, excluding this participant shifts the mean per-trial ELPD by less than 5% (0.713 full sample vs. 0.677 excluding Participant 5), indicating their influence on the overall predictive fit estimate is limited. Together, the scarcity of high-$k$ trials and the absence of disproportionate participant-level leverage support the robustness of model evaluation to undue influence from individual observations or participants.

Table S2: Participant-level aggregates of pointwise PSIS-LOO quantities. For each participant, we report the sum of pointwise expected log predictive density ($\text{elpd}_{\text{loo}}$) and the sum of effective number of parameters ($p_{\text{loo}}$) across that participant's trials.

| Participant | $\text{elpd}_{\text{loo}}$ | $p_{\text{loo}}$ |
|---:|---:|---:|
| 5 | −387 | 23.40 |
| 3 | −329 | 12.40 |
| 7 | −306 | 10.90 |
| 1 | −304 | 11.10 |
| 12 | −274 | 11.00 |
| 13 | −268 | 10.70 |
| 6 | −244 | 12.90 |
| 9 | −237 | 10.20 |
| 4 | −236 | 9.77 |
| 10 | −223 | 9.79 |
| 11 | −198 | 11.10 |
| 8 | −185 | 11.40 |
| 2 | −160 | 9.78 |
| 14 | −145 | 12.80 |

### S3 Posterior Predictive Checks

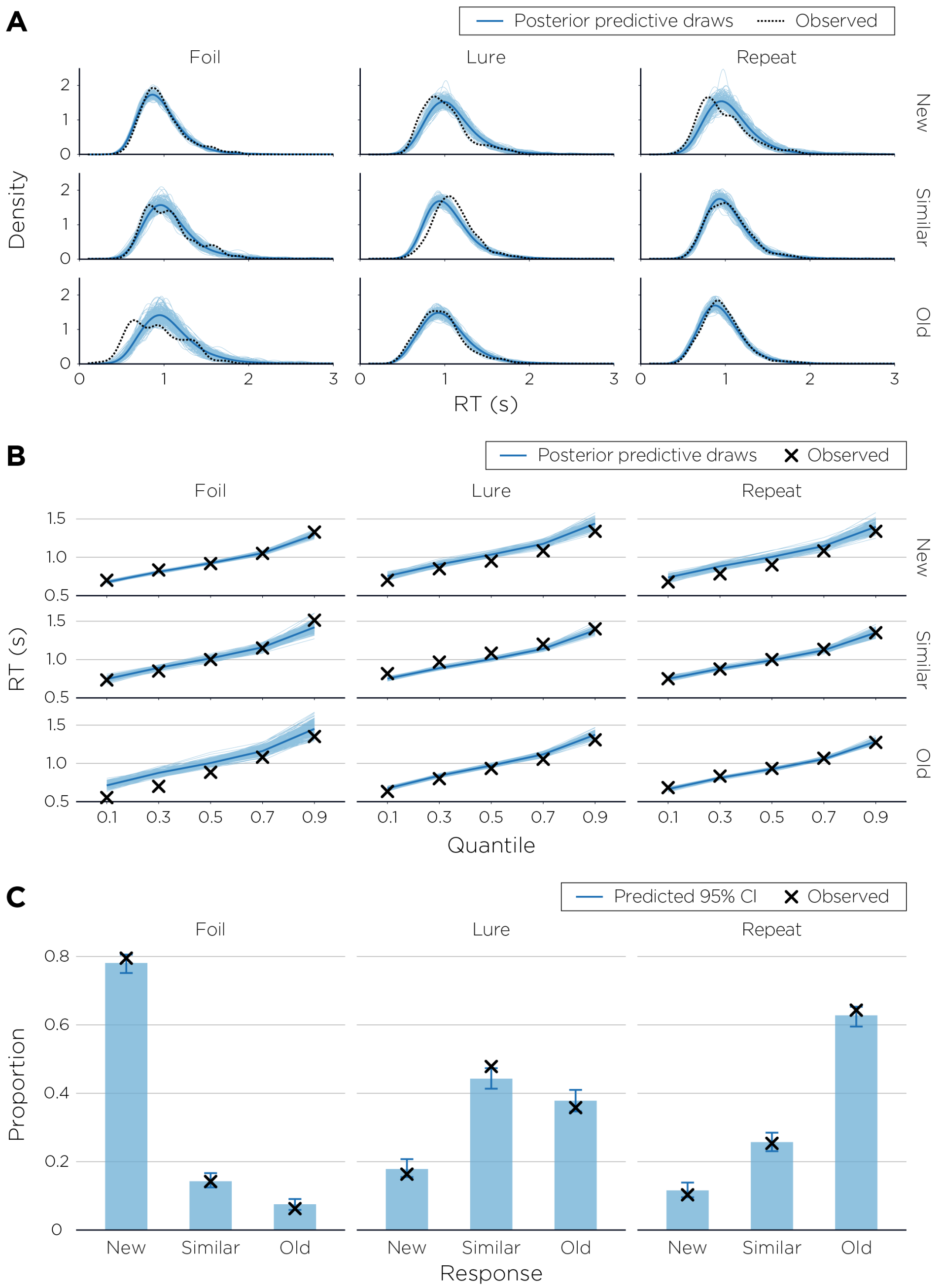


Figure S3: Posterior predictive checks for the LBA model. Posterior predictive distributions (blue) are compared against observed data (black) across three stimulus types (Repeat, Lure, Foil) and three response categories ("Old", "Similar", "New"). (A) RT density distributions. Thin blue lines represent 200 individual posterior predictive draws; the bold blue line shows the mean predicted density; the dashed black line shows the observed RT distribution. (B) RT quantile plots across the 10th, 30th, 50th, 70th, and 90th percentiles. Blue lines show posterior predictive draws and the bold line the median predicted quantile; crosses (×) indicate observed quantiles. (C) Choice proportion plots. Bars represent the median posterior predicted response proportion; error bars denote 95% credible intervals; crosses indicate observed proportions. The model accurately recovers the pattern of response tendencies across stimulus types.

### S4 Threshold Model and Starting Point Model

#### S4.1 Model Setup

To assess whether the observed theta–drift relationships were specific to the drift-rate locus of the LBA, or could equivalently be attributed to other decision parameters, we fitted two additional model variants. In the **threshold model**, trial-level theta power modulated the decision threshold via a log-linear link:

$$b_i = b_j \cdot \exp\bigl(\gamma_{\text{left}}[s]\,\theta_{\text{left}} + \gamma_{\text{right}}[s]\,\theta_{\text{right}}\bigr), \tag{1}$$

where $\gamma_{\text{left}}[s]$ and $\gamma_{\text{right}}[s]$ are stimulus-specific coefficients quantifying the influence of left and right hippocampal theta on the threshold, and the exponential link preserves the constraint $b > A$ across all trials. In the **starting-point model**, theta modulated the width of the starting-point distribution:

$$A_i = A_j \cdot \exp\bigl(\delta_{\text{left}}[s]\,\theta_{\text{left}} + \delta_{\text{right}}[s]\,\theta_{\text{right}}\bigr), \tag{2}$$

where $\delta_{\text{left}}[s]$ and $\delta_{\text{right}}[s]$ are the corresponding stimulus-specific coefficients. In both models, all remaining parameters (drift rates, non-decision time, drift rate variability) retained the same hierarchical structure and priors as the primary drift-rate model (see Section 2.9 of the main text). Theta regression coefficients ($\gamma$, $\delta$) received $\mathcal{N}(0, 0.5)$ priors, identical to the $\beta$ priors in the drift-rate model, ensuring comparability. Both alternative models were estimated using the same HMC sampling settings as the primary model (4 chains, 5,000 iterations, 1,000 warmup, `adapt_delta`$= 0.95$, `max_treedepth`$= 12$).

Model comparison was performed using leave-one-out cross-validation (LOO-CV) as implemented in the **loo** package in R, with relative efficiency corrections for MCMC autocorrelation applied prior to computing LOO estimates. The expected log pointwise predictive density ($\text{elpd}_{\text{loo}}$) and its standard error were used as the basis for comparison; differences in ELPD ($\Delta$ELPD) were interpreted as meaningful only when $|\Delta\text{ELPD}/\text{SE}_{\text{diff}}| > 2$.

#### S4.2 Model Comparison

LOO-CV diagnostics were excellent across all three models. In each case, $\geq 99.96\%$ of trials received good Pareto-$k$ values ($k \leq 0.7$), with only two trials flagged per model ($< 0.05\%$ of trials), and no trials with very bad values ($k > 1$) in any model. Inspection confirmed that the same two trials were flagged across all three models, indicating that these observations reflect data characteristics rather than model-specific instability.

ELPD estimates and model ranks are presented in Table S3. The starting-point model achieved the numerically highest ELPD ($-3496.1$, SE $= 82.3$), followed closely by the threshold model ($\Delta$ELPD $= -0.3$, $\text{SE}_{\text{diff}} = 1.7$) and the drift-rate model ($\Delta$ELPD $= -2.3$, $\text{SE}_{\text{diff}} = 6.0$). All pairwise $\Delta$ELPD-to-SE ratios were far below 2 (ratios: 0.18 and 0.38, respectively), indicating that no model was distinguishable from the others in out-of-sample predictive accuracy. Accordingly, the three models cannot be ranked on predictive grounds alone.

Table S3: LOO-CV model comparison for three LBA variants differing in the locus of theta modulation.

| Model | $\text{elpd}_{\text{loo}}$ | SE | $p_{\text{loo}}$ | $\Delta$ELPD |
|---|---|---|---|---|
| Theta → Starting Point | −3496.1 | 82.3 | 153.6 | 0.0 |
| Theta → Threshold | −3496.4 | 82.6 | 152.6 | −0.3 |
| Theta → Drift Rate | −3498.4 | 82.8 | 167.1 | −2.3 |

$\text{elpd}_{\text{loo}}$ = expected log pointwise predictive density; SE = standard error of $\text{elpd}_{\text{loo}}$; $p_{\text{loo}}$ = effective number of parameters; $\Delta$ELPD = difference in ELPD relative to the best-fitting model (Starting Point). All models had $\leq 2$ trials with Pareto-$k > 0.7$ ($< 0.05\%$ of trials). No pairwise $\Delta\text{ELPD}/\text{SE}_{\text{diff}}$ ratio exceeded 0.4, indicating no model was distinguishable on predictive grounds.

Despite equivalent predictive performance, the theta coefficients within the threshold and starting-point models were uninformative. In the threshold model, all six $\gamma$ coefficients (3 stimulus types $\times$ 2 hemispheres) were negligible in magnitude and spanned zero: Foil ($\gamma_{\text{left}} = 0.005$ [−0.005, 0.015], $\gamma_{\text{right}} = -0.005$ [−0.016, 0.005]); Lure ($\gamma_{\text{left}} = 0.006$ [−0.003, 0.016], $\gamma_{\text{right}} = 0.001$ [−0.009, 0.010]); Repeat ($\gamma_{\text{left}} = 0.000$ [−0.009, 0.010], $\gamma_{\text{right}} = 0.003$ [−0.007, 0.013]). Similarly, all six $\delta$ coefficients in the starting-point model were near-zero with wide, zero-spanning credible intervals: Foil ($\delta_{\text{left}} = -0.012$ [−0.052, 0.024], $\delta_{\text{right}} = 0.025$ [−0.011, 0.061]); Lure ($\delta_{\text{left}} = -0.025$ [−0.060, 0.009], $\delta_{\text{right}} = 0.000$ [−0.032, 0.033]); Repeat ($\delta_{\text{left}} = 0.003$ [−0.029, 0.035], $\delta_{\text{right}} = -0.016$ [−0.050, 0.018]). All intervals reported as 95% credible intervals.

All three models achieved essentially equivalent predictive accuracy, but only the drift-rate model showed selective theta modulation. In contrast, theta effects on decision threshold and starting-point range were consistently near zero, with credible intervals spanning zero (Figure S4). This dissociation suggests that the theta–behavior relationship is preferentially expressed through the rate of evidence accumulation rather than through boundary setting or pre-decisional response bias. Notably, the drift-rate model used a richer parameterization (response- and stimulus-specific $\beta$ coefficients; $p_{\text{loo}} = 167.1$) than the threshold and starting-point models ($p_{\text{loo}} = 152.6$ and 153.6), yet it did not yield superior predictive accuracy. Greater flexibility without improved out-of-sample fit argues against overfitting and points to drift rate as the structural locus for condition-specific theta effects. Taken together, these findings are consistent with hippocampal theta power modulating evidence accumulation, although predictive model comparisons alone cannot uniquely localize the underlying mechanistic effect.

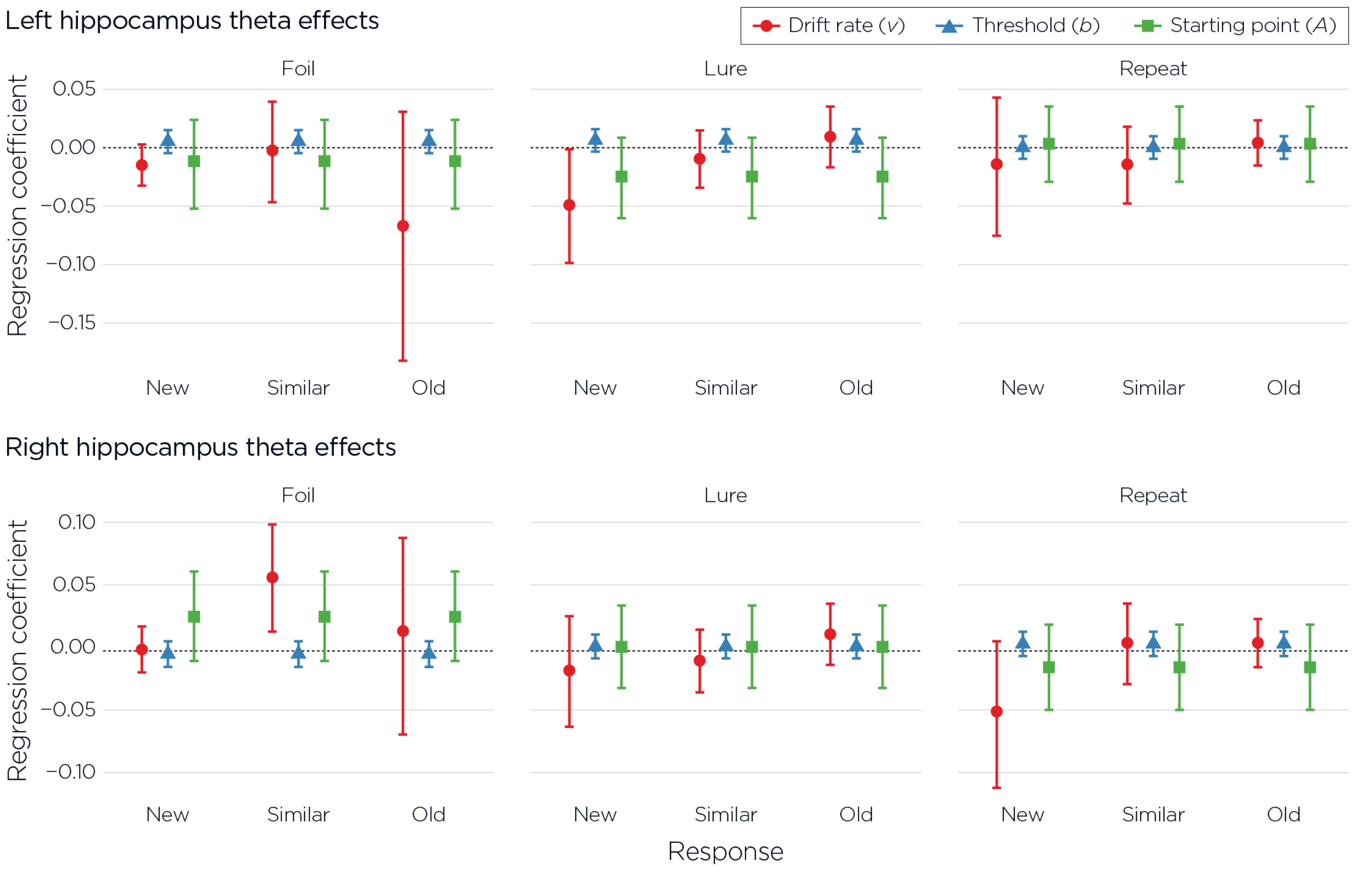


Figure S4: Posterior mean regression coefficients (±95% credible intervals) quantifying the association between hippocampal theta power and LBA decision parameters are shown separately for the left (top row) and right (bottom row) hippocampus. Effects are displayed for each stimulus type (Foil, Lure, Repeat) and response category (New, Similar, Old). Red circles indicate drift rate ($v$) coefficients from the primary drift-rate model; blue triangles indicate decision threshold ($b$) coefficients from the threshold model; green squares indicate starting-point ($A$) coefficients from the starting-point model. The horizontal dashed line marks zero. Drift rate coefficients show stimulus- and response-specific modulation by theta power, whereas threshold and starting-point coefficients consistently center near zero with credible intervals spanning zero, indicating no reliable influence of hippocampal theta on decision criterion or initial response bias.
